# Leveraging Deep Learning to Enhance MRI for Brain Disorders

**DOI:** 10.1101/2025.02.10.25321126

**Authors:** Yuxiang Dai, Renat Yakupov, Yunman Xia, Wei Tang, Tristram Lett, Frauke Nees, Elli Polemiti, Jean-Charles Roy, Ying-Hua Chu, Nilakshi Vaidya, Chengyan Wang, Xiao Chang, Chao Xie, Boyu Zhang, Xingzhong Zhao, Rencheng Zheng, Liping Zheng, Shuai Xu, Sinead King, Yuning Zhang, Zuo Zhang, Arun L Bokde, Argyris Stringaris, Dimitri Papadopoulos, Gareth J Barker, Hervé Lemaître, Hedi Kebir, Henrik Walter, Julia Sinclair, Rüdiger Brühl, Robert Whelan, Ulrike Schmidt, Jianfeng Feng, Sylvane Desrivières, Tianye Jia, Emrah Duezel, Andre Marquand, He Wang, Gunter Schumann

## Abstract

The limited availability and high cost of 7 Tesla (7T) structural MRI hinder its widespread application despite its superior imaging quality. This study introduces a High Frequency-Generative Adversarial Network (HF-GAN) to predict three-dimensional 7T-equivalent (P7T) images from standard 3T structural MRI scans, offering a cost-effective alternative. HF-GAN was trained on paired 3T and 7T MRI data and validated on external datasets, including STRATIFY/ESTRA (N=671) and ADNI2 (N=643), covering psychiatric and neurodegenerative disorders. Results indicate that P7T images generally exhibit enhanced contrast and preservation of fine structural details comparable to 7T and better than 3T, including improved sensitivity in detecting disease-related differences in key brain regions such as the thalamus, caudate, putamen, and frontal cortical areas. The partial η^2^ values revealed that P7T explained a higher proportion of variance compared to 3T in several comparisons, highlighting its improved sensitivity to disease-related structural changes. These findings demonstrate that HF-GAN effectively enhances 3T MRI data quality, providing a scalable solution for research and clinical applications in neurodegenerative and psychiatric disorders. Additional validations in brain and other organ systems are warranted to further advance clinical translation.

## Introduction

Neuroimaging contributes to the detection and characterization of brain disorders by identifying structural and functional alterations associated with these conditions. Over recent years, neuroimaging techniques have seen significant refinement; however, they still fall short of delivering the accuracy needed for timely diagnosis and effective treatment, particularly in psychiatric and neurodegenerative disorders [1–4]. Current imaging techniques, such as 3 Tesla (3T) magnetic resonance imaging (MRI), are often inadequate in capturing the brain structural variations that underlie these conditions. These limitations stem primarily from insufficient sensitivity to detect subtle variations in psychiatric disorders or to fully characterize extensive regional changes associated with neurodegenerative diseases. While biological heterogeneity contributes to the complexity of brain structural variations, the limitations of current imaging techniques are also a significant factor, particularly in their sensitivity to subtle changes and their ability to capture extensive regional alterations. This constraint can contribute to delays in diagnosis and limit the precision of treatment strategies. [5–7].

The most advanced technology for human *in vivo* structural neuroimaging is 7 Tesla (7T) MRI, which offers enhanced resolution and contrast, making it particularly effective in differentiating structures within specific brain regions [8,9]. The higher field strength allows for the detection of microstructural changes that are often missed by 3T MRI [10–12]. This makes 7T very useful in the study of neurodegenerative and psychiatric disorders [13–15]. However, the use of 7T MRI remains limited due to its high cost, limited availability, noisy image acquisition and the logistical challenges associated with its implementation [16,17]. Furthermore, large-scale studies involving 7T MRI are rare, as acquiring sufficiently large sample sizes to reliably demonstrate and quantify diagnostic effects is difficult under these constraints [18].

To address this dilemma, artificial intelligence (AI) was explored as a solution to predict 7T-equivalent images from existing structural 3T MRI scans. While prior methods for image prediction and enhancement have been developed [19–25], they face several limitations. Specifically, many rely on 2D slice-based predictions, which fail to preserve the spatial continuity of 3D brain structures [19,20,23–25]. Others emphasize contrast enhancement but often lack the ability to capture fine-grained diagnostic details [21,22]. Furthermore, most existing approaches are trained on single datasets, raising concerns about their generalizability across diverse populations and imaging protocols [19–23]. Finally, these methods rarely assess the clinical relevance of the predicted 7T images, leaving their practical utility for diagnosis and treatment unclear.

To overcome these limitations, we developed a novel 3T to 7T prediction model (3D) based on High Frequency-Generative Adversarial Network (HF-GAN). We trained and validated this model using two independent datasets with real 7T data, allowing us to develop and fine-tune the method. We then applied the model to two additional clinical datasets (STRATIFY/ESTRA and ADNI2), comprising patients with Neurodegeneration [26–29], Alcohol Use Disorder [30,31], Major Depression Disorder [32,33] and Eating Disorders [34,35].

Our aim is to enhance the utility of structural 3T imaging by developing a scalable method for generating P7T images, enabling more precise characterization of brain structure using existing large 3T datasets and facilitating earlier detection of brain disorders.

## Results

### Summary of major analytic steps (Figure 1)

To predict 3D 7T brain scans from 3T data (Figure 1a), we first trained the High Frequency-Generative Adversarial Network (HF-GAN) on 30 young adults from Magdeburg, Germany, scanned with both 3T and 7T machines (see Methods). We then trained HF-GAN on an independent dataset of 43 young adults from Shanghai, China for external validation, scanned with both 3T and 7T (see Methods and Extended Table 1). We used five-fold cross-validation to obtain all predicted 7T (P7T) T1 images for each dataset. Next (Figure 1b), we segmented the original and predicted MRIs, using Voxel-Based Morphometry (VBM) to compare gray matter volume in 3T vs. 7T and P7T vs. 7T (European brains template for Magdeburg data and East Asian brains template for Shanghai data). To identify enhanced brain regions, we relied on widely adopted metrics like cortical thickness and subcortical volume. Recognizing HF-GAN’s effect on image contrast, we also included gray-white matter contrast (GWR) (see Methods) to better capture variations in regional brain features based on Freesurfer segmentation (Figure 1c).

**Figure 1:**
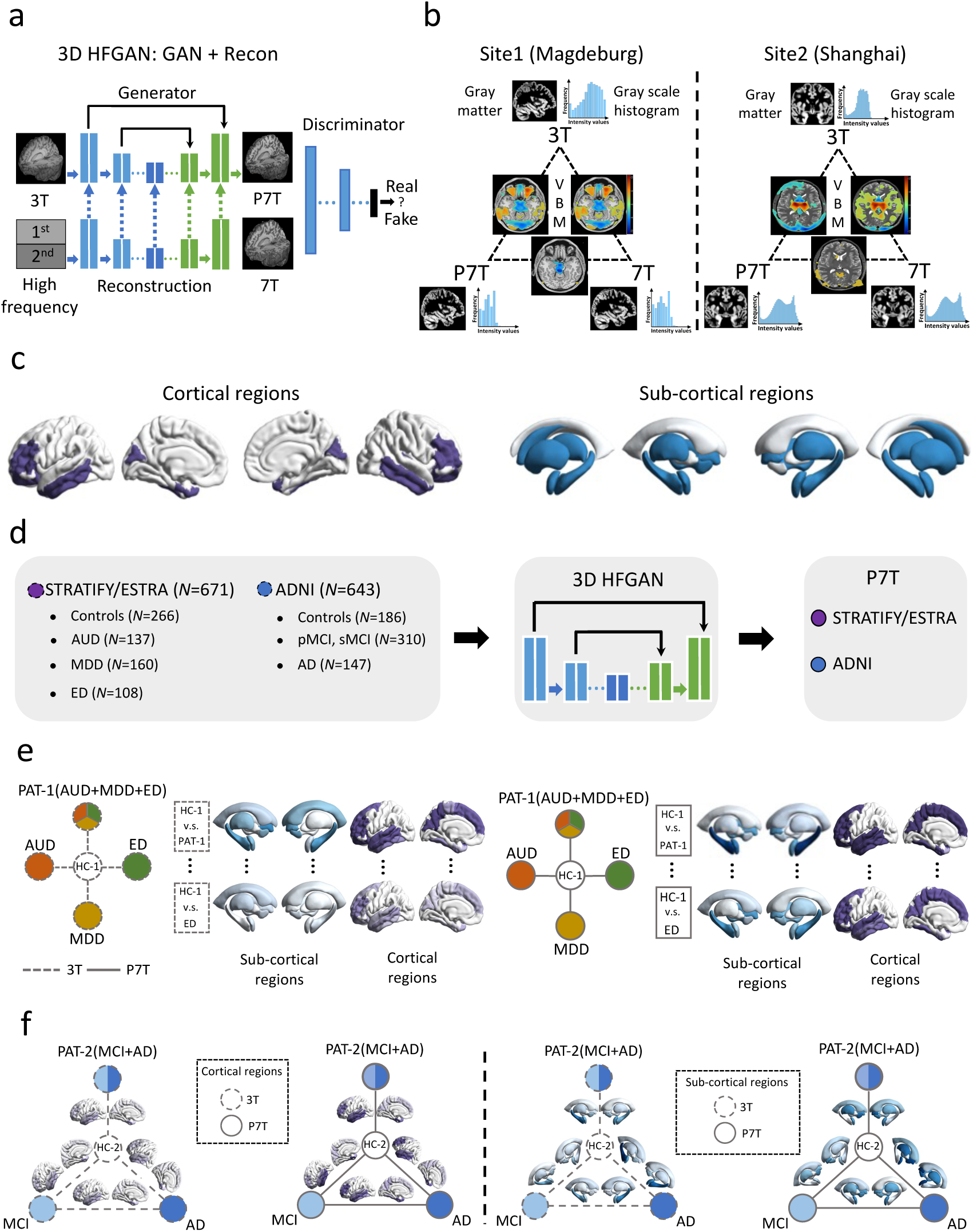
Characterization of the study design. a We established a 3D model called HFGAN using the preprocessed dataset. b This study primarily focused on validating the effectiveness of P7T in enhancing contrast and image details for entire brain. c In specific cortical and subcortical brain areas, we conducted statistical analyses to investigate the differences in structural indicators among 3T, P7T, and 7T. d We extended our method to two clinical cohorts for respective P7T. e We applied our methodology to the STRATIFY/ESTRA cohort to explore differential alterations between the 3T to P7T transition in the healthy group and the disease control groups. AUD, alcohol use disorder; MDD, major depressive disorder; ED, eating disorder. f We verified our method in older adults with neurodegenerative conditions of the ADNI cohort for case-control comparisons. MCI, Mild Cognitive Impairment; pMCI, Progressive Mild Cognitive Impairment; sMCI, Stable Mild Cognitive Impairment; AD, Alzheimer’s.

We hypothesized that a more detailed depiction of cortical and subcortical regions with P7T MRI will identify brain regions involved in mental illness that may be absent from lower contrast 3T scans. We therefore applied our model in young adults with major depressive disorder (MDD) (n=169), alcohol use disorder (AUD) (n=137), eating disorders (ED) (n=108) and healthy controls (HC-1) (n=266) (STRATIFY/ESTRA study) (Figure 1e) as well as in older adults with mild cognitive impairment (MCI) (n=310), Alzheimer’s disease (AD) (n=147) and healthy controls (HC-2) (n=186) (ADNI2 cohort) (Figure 1f). We then conducted case-control comparisons within each cohort to identify brain structural alterations associated with these disorders. Specifically, we compared each patient group (MDD, AUD, and ED in STRATIFY/ESTRA; MCI and AD in ADNI2) to their respective healthy control groups (HC-1 and HC-2) to examine disease-related changes in brain regions.

### Whole brain visual inspection

Our method statically improves the differences in contrast in P7T images, closely resembling 7T MRI quality. As seen in Figure 2a, the delineation of gray and white matter, particularly in cortical regions like the cingulate gyrus, is clearer, and the structural complexity matches that of 7T images. P7T also captures vascular details within the insular cortex and shows marked enhancements in the caudate and putamen. The grayscale histogram of P7T is comparable to the distribution seen in 7T MRIs. In gray matter structures shown in Figure 2b, the P7T images are not blurred but reveal intricate internal details, underscoring our method’s ability to enhance contrast and detail fidelity.

**Figure 2:**
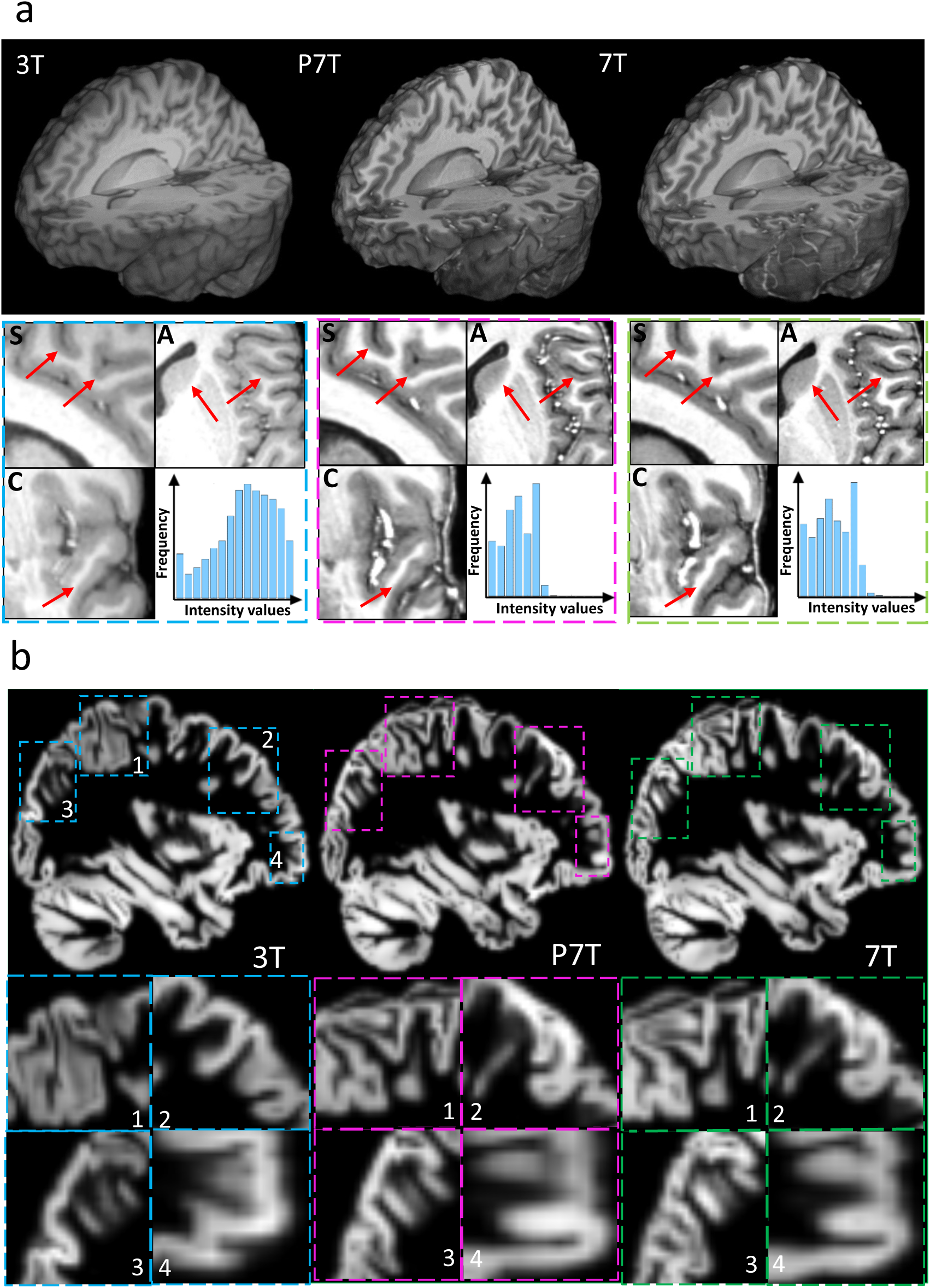
The comparisons at the imaging level of Magdeburg dataset. a A testing case for 3T, P7T prediction and 7T which have been showcased in 3D view, along with the corresponding local regions in sagittal (S), axial (A) and coronal (C) views. The gray-level histogram distribution of this testing data has also been calculated and displayed. The X-axis is intensity value and the Y-axis is the number of voxels (Frequency). Among them, the blue box is 3T, the purple box is P7T prediction, and the green box is 7T. b We demonstrated the gray matter segmentation of the testing case.

We tested our method on the Shanghai dataset, which contains T1 MP2RAGE scans and has different contrast than the Magdeburg T1 MPRAGE. P7T images exhibited enhanced gray-white matter boundary delineation (see arrow in Figure 2a), and the histogram confirmed the P7T images’ similarity to actual 7T scans. In the zoomed areas of 3T sagittal view (Suppl. Figure 3a), there were artifacts near the hippocampus and striatum, which might affect tissue segmentation accuracy. These artifacts were not present in the P7T and 7T images.

In addition, we analyzed statistical differences in gray matter volume using VBM for the Magdeburg and Shanghai samples with CAT12[36]. The differences between 3T and 7T scans were generally larger in terms of T-values compared to those between P7T and real 7T scans, particularly in areas like the central sulcus and hippocampus. The T-values for 3T vs. 7T were higher than those for P7T vs. 7T in these regions. In contrast, compared to 3T, the P7T showed only minor differences in regions like the precuneus, middle temporal gyrus, and inferior temporal gyrus (Suppl Figure 4).

### Whole brain quantitative assessments

To assess contrast enhancement, we used Contrast to Noise Ratio (CNR) and Signal to Noise Ratio (SNR) metrics (Suppl. Figure 5 and Method). We focused on the hippocampus in the Magdeburg dataset as manual segmentation data were available of this region [37]. The CNR and SNR values of 7T predictions were between those of 3T and 7T, leaning closer to 7T but not exceeding it (Suppl. Figure 5). This indicates that HFGAN’s 7T predictions enhance contrast appropriately without overly increasing tissue contrast.

Whole-brain analysis of 3T and P7T groups, including imaging and quantitative metrics (Extended Table 2), shows that 7T predictions significantly improve image quality. This enhancement is evident in better image contrast (PSNR in Extended Table 2) and finer detail (SSIM in Extended Table2). Following these results, we further explored the impact of 7T predictions on additional brain regions using automatic segmentation.

### Regional quantitative assessments

We first evaluated performance of different automatic segmentation programs in the Magdeburg dataset, using manual segmentation of the 7T hippocampus as a reference. Dice coefficients were calculated for 3T and P7T hippocampus to compare among segmentation programs including: Freesurfer [38], ANTs [39], and FSL [40]. Freesurfer had higher Dice coefficients for segmenting both left (0.37) and right (0.36) hippocampus in P7T images than the other tools (ANTs: left-0.36, right-0.35; FSL: left-0.27, right-0.29) (Suppl. Figure 6). Additionally, Freesurfer’s segmentation of P7T images significantly differed from 3T (left: FDR<0.0001; right: FDR<0.01), especially in the right hippocampus, where FSL showed greater disparity but lower overall Dice values. Therefore, we chose Freesurfer for its superior accuracy in P7T segmentation of our datasets for detailed quantitative brain region analyses.

### Sub-cortical quantitative assessments

We utilized Freesurfer’s Automatic Subcortical Segmentation Template (ASEG) to analyze the volume of subcortical structures. We captured the resolution within each region by quantitatively measuring signal contrast at the regional level. There were no significant differences (FDR>0.05) between P7T and 7T, but notable differences (FDR<0.05) between 3T and both P7T and 7T GWR and also volume, indicating HFGAN’s effective enhancement (Figure 3a). We found significant subcortical enhancement, particularly in the thalamus, amygdala, caudate nucleus, and hippocampus (Figure 3a). The detailed results of GWR and volume can be seen in Suppl. Figure 7. Dice coefficients for 3T, P7T, and ground truth 7T were calculated for brain areas (Figure 3d) showing significantly improved segmentation in P7T compared to 3T (Suppl. Figure 8a).

**Figure 3:**
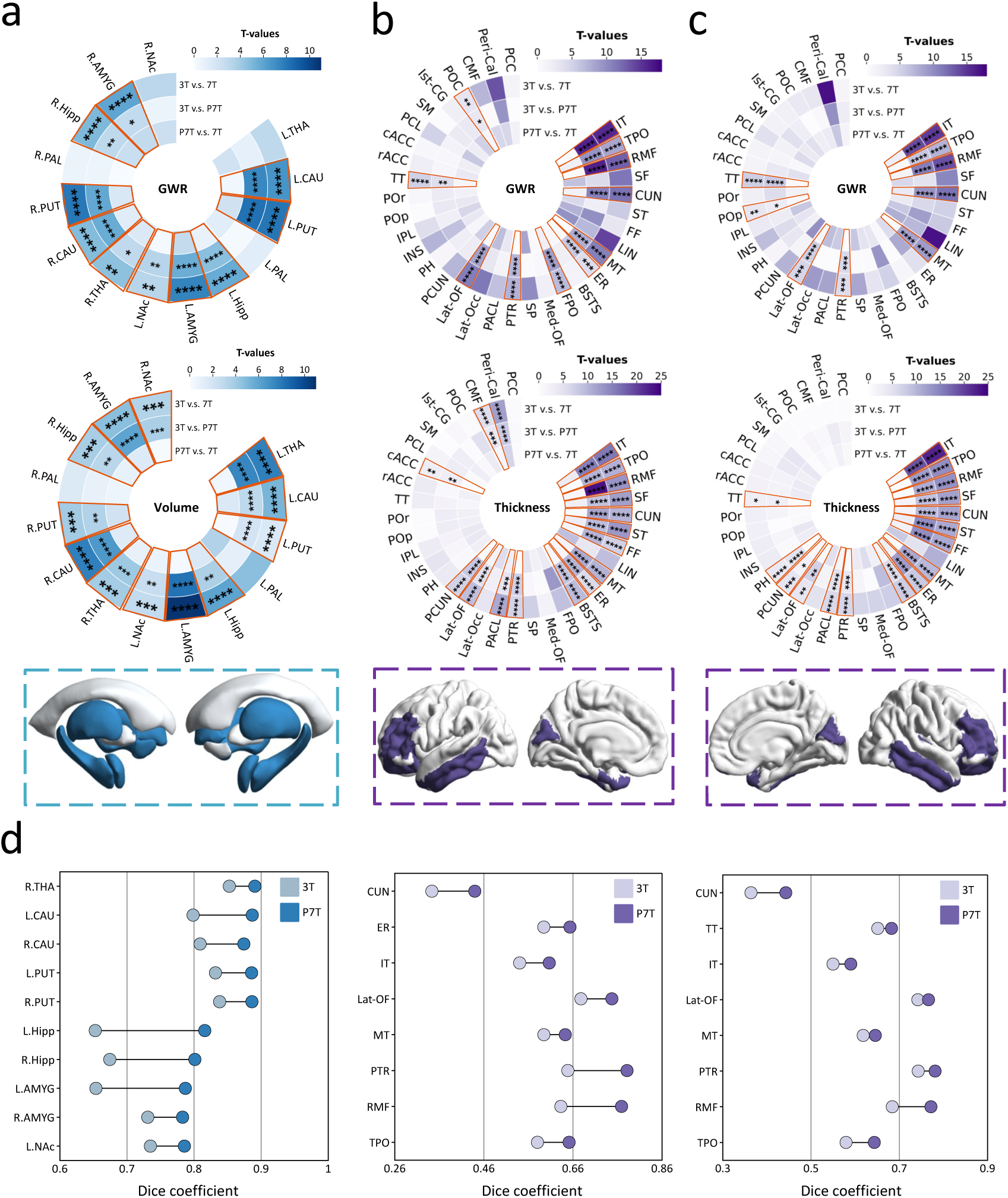
The significantly enhanced brain regions by HFGAN and segmentation Dice results of these regions. We first performed a paired t-tests on the data groups based on all the sample data. The statistical T-values were then mapped onto the ASEG and Desikan-Killiany (DK) templates, enabling us to observe the spatial distribution of differences in the brain. We calculated the segmentation Dice coefficient for 3T and P7T using 7T as the gold standard, then demonstrated the 3T and P7T dice on the dumbbell figures. a The group differences in all subcortical brain regions based on GWR and volume. b The group differences in left hemisphere based on GWR and thickness. c The group differences in right hemisphere based on GWR and thickness. d The calculated dice coefficients for 3T and P7T based on significantly enhanced sub-cortical and cortical regions shown by dumbbell figures. The x-axis represents the dice coefficient. From left to right, they are the subcortical brain regions, the left hemispheres, and the right hemispheres. ns: no significance; *: FDR < 0.05; **: FDR < 0.05; ***: FDR < 0.001; ****: FDR < 0.0001. The full name of the abbreviated brain regions can be found in Supplemental Table 3.

### Cortical quantitative assessments

Significant differences in cortical contrast and thickness between 3T and P7T were seen in areas of the temporal, frontal, occipital, and limbic lobes in both hemispheres. In particular, the temporal pole, the pars triangularis and rostral middle frontal cortex, as well as the occipital cuneus showed marked differences (FDR<0.05) when comparing 3T and P7T, as well as 3T and 7T. No differences were observed between P7T and 7T. (Figures 3b and 3c). Other cortical structures showed similar thickness as 3T, indicating no significant enhancement.

Both P7T and 7T scans generally showed consistent changes compared to 3T across all brain regions (Suppl. Figure 8). Similar to 7T scans [41], P7T did not uniformly reduce thickness measures across cortical regions but showed region-specific variations. For instance, in the frontal structures, the lateral orbitofrontal areas had reduced thickness in 7T and P7T compared to 3T, while areas like pars triangularis and the rostral middle frontal lobe showed increased thickness. A similar differentiated pattern was observed in the cuneus lobe, indicating the fidelity and precision of our prediction method.

Figure 3d demonstrates that P7T exhibits segmentation performance closer to 7T compared to 3T. Suppl. Figures 8b and 8c show individual case segmentations and highlight significant improvements (FDR<0.05) in segmentation accuracy for P7T images across all enhanced brain areas.

### P7T application in patients with psychiatric and neurodegenerative disorders and controls

We applied HFGAN to the STRATIFY/ESTRA [42] and ADNI2 [43] datasets, with visualization results shown in Suppl. Figures 10 and 13. Notably, P7T images exhibit higher contrast and more detailed features across both datasets (see arrows in Suppl. Figures 10 and 13). We hypothesized that prediction of 7T images will reveal correlations of neurodegenerative and psychiatric disorders with brain structure that are not detectable in 3T images. We therefore applied HF-GAN in the STRATIFY/ESTRA sample of patients with AUD, ED, MDD and healthy controls (HC-1) (Extended Table 3). We also applied HF-GAN to the ADNI2 sample of patients with MCI including stable MCI (sMCI) and progressive MCI (pMCI), AD and healthy controls (HC-2) (Extended Table 4). We then compared significant differences between cases and controls in all brain regions (Figures 4, 5 and Suppl. Tables 6, 10). In suppl. Figures 9 and 12, we also present results of the comparison of enhanced cortical/sub-cortical regions only. The proportion of explained variance is indicated in partial η^2^ values (Suppl. Figures 12 and 19). The details of the partial η^2^ analysis is in Statistical analysis of the Method section.

**Figure 4:**
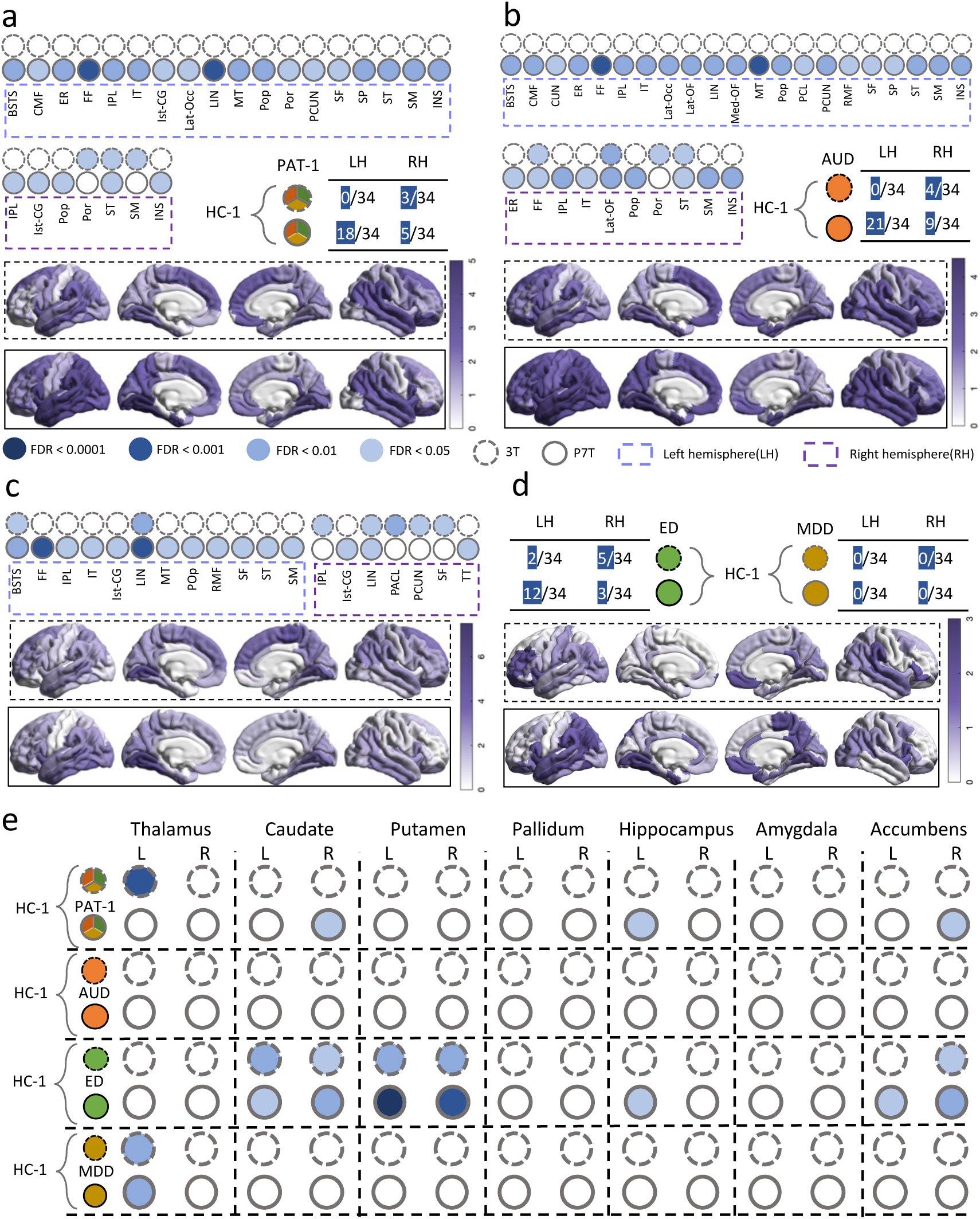
The application of STRATIFY/ESTRA. We used a generalized linear model (GLM) to compare cortical thickness or gray-white matter ratio (GWR) among several disorder groups (AUD, ED and MDD) and healthy controls (HC-1), adjusting for factors like age, gender, and scanning sites. For cortical thickness, we present the significant FDR levels of brain regions, the proportion of brain regions showing significant differences (For example, 18/34 indicates that 18 out of 34 brain regions exhibit significant differences based on FDR correction), and the absolute T-values mapped onto the brain. For sub-cortical GWR, we present the significant FDR levels. a Cortical comparison between HC-1 and PAT-1. b Cortical comparison between HC-1 and AUD. c Cortical comparison between HC-1 and ED. d Cortical comparison between HC-1 and MDD (MDD has no regions with significant difference). e Sub-cortical comparison across all disorder groups. The results of cortical GWR and sub-cortical volume can be seen in Supplemental Figure 11.

### Alcohol Use Disorder, Major Depression Disorder and Eating Disorders (STRATIFY/ESTRA)

We applied HF-GAN to the STRATIFY/ESTRA sample of young adult patients with non-neurodegenerative psychiatric disorders. When comparing all patients (PAT-1) to healthy controls (HC-1), we found in P7T significant differences (FDR <0.05) in thickness in cortical (18/34 regions, left; 5/34 regions, right) and contrast in subcortical (3/14) regions, whereas in 3T there were few significant associations with cortical brain regions (3/34, right) and 1/34 subcortical associations.

When investigating individual disorders, we found that AUD vs. controls in P7T showed significant differences in contrast in several temporal brain regions, as well as in the frontal and occipital cortex (21/34, left; 9/34 right) (Figure. 4; Suppl. Table S6). 3T showed fewer cortical areas with significant differences (0/34, left; 4/34, right) (Figure. 4b; Suppl. Table S6). There were no significant differences in either P7T or 3T in subcortical regions.

ED patients vs. control showed enhanced differences in P7T in both cortical and subcortical regions. Whereas 3T showed few significant cortical differences (2/34, left; 5/34, right), in P7T we found differences in the parietal, frontal and temporal cortices (12/34, left; 3/34, right). P7T (7/14 regions) was more sensitive than 3T (5/14 regions) in detecting differences in subcortical regions, adding hippocampus and di-encephalon to caudate, putamen and n. accumbens that were detected by 3T (Figure. 4c; Suppl. Table S6).

In MDD patients vs. controls applying P7T in subcortical areas yielded modest enhancement compared to 3T, namely in the Left Thalamus. No significant association was found in cortical areas with either P7T or 3T (Figure. 4d; Suppl. Table S6).

When measuring cortical contrast and subcortical volume in STRATIFY/ESTRA, we found that P7T was more sensitive to detect differences in the diseased brain regions in the left temporal lobe that were not registered with 3T (Suppl. Figures 11a-d, Suppl. Table S6). Compared to 3T, P7T was more sensitive to detect changes in the subcortical hippocampus, particular in the HC-1 versus. (v.s.) PAT-1 and HC-1 v.s. MDD comparisons, but exhibited a slight decrease in sensitivity for the pallidum and amygdala in the HC-1 v.s. PAT-1 and HC-1 v.s. AUD (Suppl. Figure 11e, Suppl. Table S6). There was asymmetry towards the left hemisphere, where correlations with disease were stronger. (Figure 4, Suppl. Figure 11).

When evaluating the explained variance of psychiatric disorders detection across the whole brain with P7T, we found that P7T generally exhibited lower partial η^2^values in the cortical GWR in patients compared to 3T, but higher partial η^2^ values for cortical thickness across the whole brain (see Suppl. Figure 12a). In the subcortical structures of the whole brain, except for the volume in the PAT-1 and ED groups, P7T showed a higher proportion of explained variance in GWR and volume in all other groups compared to 3T (see Suppl. Figure 12b).

### Mild Cognitive Impairment and Alzheimer Dementia (ADNI2)

We then applied HF-GAN to the ADNI2 samples of patients with neurodegenerative disorders. In comparisons of regional subcortical GWR in ADNI2 of all patients vs. controls, P7T was more sensitive than 3T (significant association in 10/14 regions in P7T compared to 4/14 regions in 3T). While this advantage was observed in all subgroups (Figure 5a and 5b), it was particularly prominent in the early stages of disease, when comparing MCI vs. controls. Here P7T detected 8/14 regions, compared to 4/14 in 3T. P7T was also more sensitive than 3T in detecting differences between MCI and AD with 10/14 regions vs. 3/14, respectively. Whereas 3T only detected contrast differences in hippocampus and amygdala, P7T detected additional significant differences in caudate, putamen, n. accumbens and thalamus.

**Figure 5:**
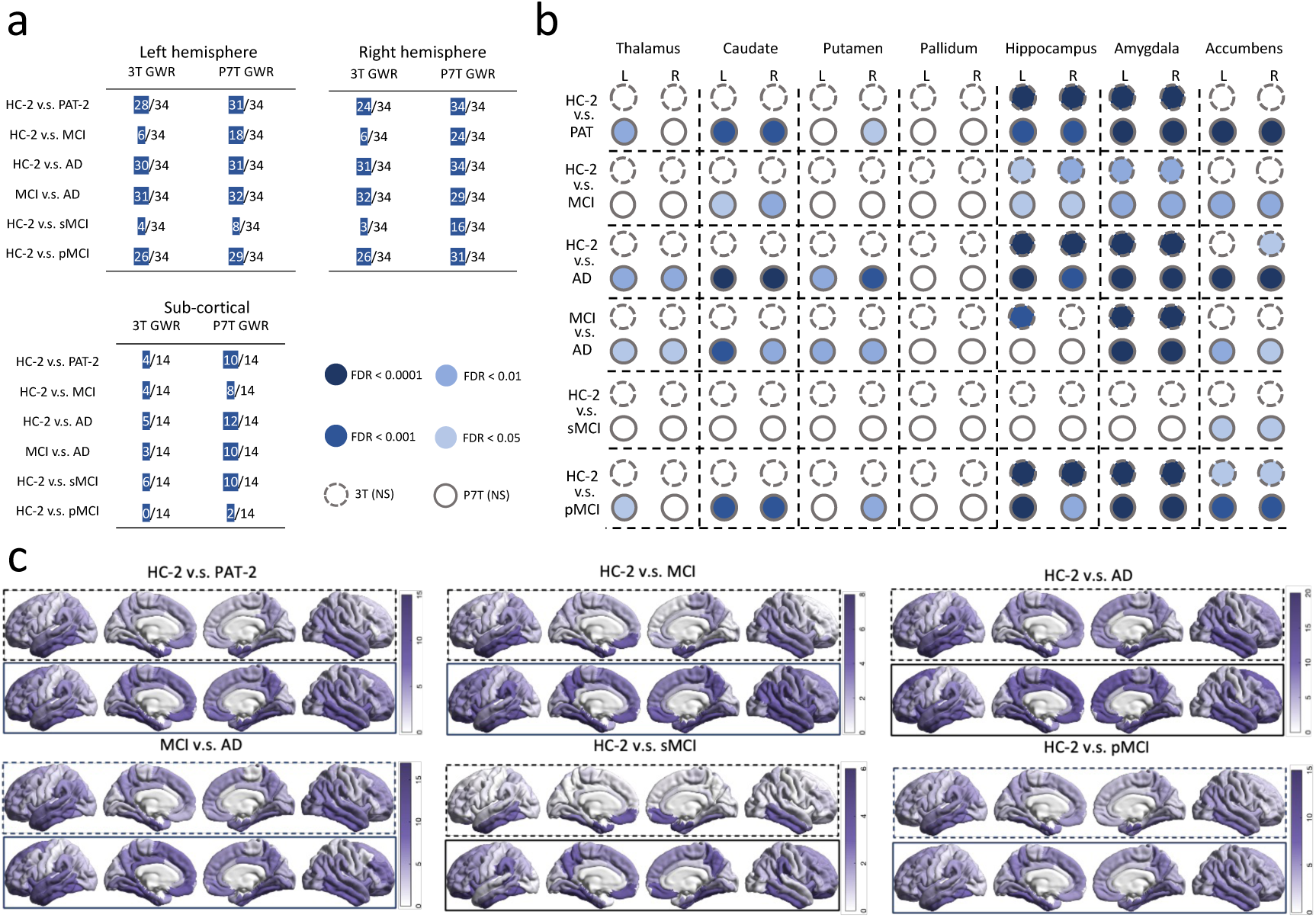
The application of ADNI. We examined differences between diseases (AD, MCI composed of sMCI and pMCI) and control groups (HC-2) in significantly affected brain areas. By measuring brain region GWR, we used GLM to adjust for factors like age, weights and gender, enabling us to compare regional differences across the groups. a We show the number of cortical regions with significant differences for case-control comparisons. b We show significant FDR differences in sub-cortical regions of GWR. c We show the proportion of cortical regions with significant differences and the statistical absolute T values mapped onto cortical regions. The results of cortical thickness and sub-cortical volume can be seen in Supplemental Figure 14.

When comparing contrast of cortical brain regions, P7T (left 18/34, right 24/34 regions) proved to be a more sensitive diagnostic tool than 3T (left 6/34, right 6/34 regions) to detect early signs of neurodegeneration in MCI, particularly in frontal cortical areas (Figure 5c, Suppl. Figure 15). When comparing established disease, P7T and 3T performed almost similar. AD vs. healthy controls in P7T showed significant associations in 33/34 (left) and 34/34 (regions) regions, and in 3T 30/34 (left) and 31/34 (right) regions (Figure 5a, Suppl. Figure 15). In the detection of sMCI, P7T demonstrated superior performance compared to 3T in both hemispheres (Left: 8/34 vs. 4/34; Right: 16/34 vs. 3/34). Similarly, in pMCI, P7T outperformed 3T with greater sensitivity (Left: 29/34 vs. 26/34; Right: 31/34 vs. 26/34). Detailed FDR and uncorrected P results can be seen in Suppl. Figures 15,19 and Suppl. Tables S8, S9. Detailed results of thickness/volume can be found in Suppl. materials.

In the partial η^2^results based on the ADNI2 data, we observed that for most comparisons - whether using GWR or thickness/volume - P7T consistently yielded higher partial η^2^ values than 3T (Suppl. Figure 19). This finding suggests that P7T offers higher proportion of explained variance or sensitivity in detecting neurodegenerative disorders-related brain alterations.

Together, these findings indicate that P7T might result in an earlier detection and more differentiated characterization of disease progression in neurodegenerative disorders (Figure 5).

## Discussion

In this study, we present a novel High Frequency-Generative Adversarial Network (HF-GAN) designed to predict 7T-equivalent images from standard 3T MRI scans, addressing key limitations in current neuroimaging practices. By incorporating high-frequency spatial details and contrast enhancement, HF-GAN surpasses existing methods that struggle to simultaneously maintain spatial continuity and preserve fine-grained anatomical details [20,21]. Our results demonstrate that P7T images generated using HF-GAN exhibit improved contrast and structural delineation compared to conventional 3T images, bringing them closer to the quality of actual 7T MRI. These findings could have profound implications for the field of neuroimaging, particularly in the early diagnosis and monitoring of neurodegenerative and psychiatric disorders.

A key finding of this study is the enhanced sensitivity of P7T in detecting disease-related brain alterations across multiple cohorts. In the STRATIFY/ESTRA dataset, which includes young adults with alcohol use disorder (AUD), major depressive disorder (MDD), and eating disorders (ED), P7T identified a greater number of significant differences in cortical and subcortical regions compared to 3T. Notably, P7T detected structural differences in regions such as the fusiform gyrus, inferior parietal cortex, and middle temporal gyrus, which were not observable with 3T. These regions have been previously implicated in AUD and ED pathology [44–46], suggesting that P7T enhances the ability to discriminate brain alterations between these disorders, providing a tool for differentiating structural changes. The potential for its use in diagnosis can be explored in future research, particularly with longitudinal data.

In the ADNI2 cohort, P7T generally demonstrated higher proportion of explained variance in distinguishing MCI and AD patients from healthy controls, with higher partial η^2^values compared to 3T across most cortical thickness and subcortical volume measurements. Notably, P7T was particularly effective in early-stage disease detection, identifying significant structural changes in the thalamus, caudate, putamen, and cortical regions within the temporal lobe [47,48], which is known to undergo early degeneration in Alzheimer’s disease. Although the frontal lobe typically shows degeneration in later stages [47,48], P7T also identified early structural changes in this region, suggesting its potential to facilitate earlier and more precise diagnosis, which is crucial for timely intervention and treatment planning.

Previous efforts to enhance 3T MRI quality using deep learning have primarily relied on 2D slice-based methods [13,17,19], which often fail to capture the 3D anatomical continuity of brain structures. Compared to these methods, HF-GAN introduces a unique advantage by leveraging high-frequency spatial information, allowing for more accurate preservation of fine details. Unlike conventional 3D CNN-based [22] approaches that primarily focus on structural boundaries and 3D GAN model [24] that is primarily designed to enhance contrast, our model optimally balances contrast enhancement and spatial coherence. The increased sensitivity and specificity observed in P7T images underscore the effectiveness of this novel approach in enhancing clinical utility.

The scalability and cost-effectiveness of HF-GAN make it an attractive alternative to 7T MRI in both clinical and research settings. Given the limited accessibility and high costs associated with 7T MRI, HF-GAN offers a feasible solution for institutions with only 3T infrastructure, enabling broader application in large-scale studies and potentially in routine clinical diagnostics. The ability to generate 7T-like images could improve patient stratification in clinical trials, enhance early-stage diagnosis, and provide a deeper understanding of disease progression in longitudinal studies [49]. Moreover, the superior contrast and structural fidelity of P7T images may facilitate the integration of neuroimaging with other modalities such as genomics and cognitive assessments, paving the way for more comprehensive investigations into the neurobiological underpinnings of brain disorders [50,51].

Despite the promising results, several limitations should be acknowledged. First, although HF-GAN demonstrated robust performance across the Magdeburg and Shanghai datasets, additional validation on multi-center cohorts with diverse scanner models and acquisition protocols (at least over 10 different scanners) is necessary to ensure generalizability. Differences in MRI acquisition parameters and demographic diversity may influence the performance of the model, and future work should focus on fine-tuning HF-GAN across broader populations.

Second, while P7T images show improved structural delineation and contrast, their direct clinical utility requires further evaluation through prospective studies. The diagnostic value of P7T-enhanced imaging in clinical decision-making, particularly in distinguishing closely related disorders such as sMCI and pMCI, should be systematically investigated. Moreover, potential artifacts introduced by the generative process must be carefully assessed to avoid misinterpretation in clinical practice.

Furthermore, while P7T images enhance structural delineation, they may underperform in certain cases. Artifacts introduced by deep learning-based enhancement could lead to false positives, requiring careful validation. Additionally, some case/control differences detected in 3T are less pronounced in P7T, possibly due to the generative model smoothing subtle variations. Further refinement of HF-GAN is needed to ensure accurate clinical interpretation.

An additional limitation observed in the STRATIFY/ESTRA cohort is hemispheric asymmetry in the number of significant brain regions detected in P7T. This asymmetry was more pronounced in GE scanner data, while no such asymmetry was observed in data from other scanners. This discrepancy may be related to scanner-specific factors, and future work could address this by training the model on GE data to improve balance across hemispheres.

Future research should also explore the integration of P7T with complementary diagnostic tools such as functional MRI and diffusion tensor imaging. Combining structural and functional data could provide a more holistic understanding of neurodegenerative and psychiatric disorders. Expanding the application of HF-GAN to pediatric populations could provide novel insights into neurodevelopmental structural changes, particularly in identifying biomarkers of pathological neurodevelopment. Additionally, the application of HF-GAN to imaging enhancement of other organ systems, such as cardiovascular, respiratory or hepatic should be explored.

This study establishes HF-GAN as a powerful tool for enhancing the quality of 3T MRI data, bridging the gap between conventional imaging techniques and ultra-high-field MRI. Our findings demonstrate the ability of P7T to achieve near 7T-level sensitivity in detecting disease-related brain changes, offering a cost-effective and scalable solution for neuroimaging research and clinical practice. With further validation, P7T could become an integral part of diagnostic workflows, enabling earlier detection and improved monitoring of brain disorders.

## Data Availability

All data produced in the present study are available upon reasonable request to the authors

## Acknowledgements

This work received support from the following sources: the European Union-funded Horizon Europe project ‘environMENTAL’ (101057429) and co-funding by UK Research and Innovation under the UK Government’s Horizon Europe funding guarantee (10041392 and 10038599) and the the National Key R&D Program of Ministry of Science and Technology of China (MOST 2023YFE0199700); the Horizon 2020-funded European Research Council Advanced Grant ‘STRATIFY’ (Brain network based stratification of reinforcement-related disorders; 695313); the German Research Foundation (COPE; 675346); the Medical Research Council and Medical Research Foundation (grants MR/R00465X/1 and MRF-058-0004-RG-DESRI: ‘ESTRA: Neurobiological underpinning of eating disorders: integrative biopsychosocial longitudinal analyses in adolescents’; MR/S020306/1 and MRF-058-0009-RG-DESR-C0759: ‘Establishing causal relationships between biopsychosocial predictors and correlates of eating disorders and their mediation by neural pathways’) and the National Institute for Health and Research (NIHR) Biomedical Research Centre (BRC) and Maudsley NHS Foundation Trust (SLaM); Medical Research Council (grant MR/W002418/1: ‘Eating Disorders: Delineating illness and recovery trajectories to inform personalized prevention and early intervention in young people (EDIFY)’); ERC Consolidator Grant (MENTALPRECISION 101001118); Medical Research Foundation (MRF-058-0014-F-ZHAN-C0866); National Key R&D Program of China (No. 2023YFF1204804), National Natural Science Foundation of China (No. 82271956, No. 62331021), Shanghai Municipal Science and Technology Explorer Project (No. 23TS1400500).

## Methods

### Datasets for training and evaluation

For network training and testing, we used two datasets with both 3T and 7T ground truth acquisition of structural brain data (Extended. Table 1). The Magdeburg dataset comprising 30 participants aged 19-35 years (mean age 25 years). Scans were conducted using Siemens Vario 3T and Siemens Magnetom 7T scanners. The Shanghai dataset involved 43 participants aged between 24 and 37 years (mean age 26 years). Scanning was carried out using Siemens Prisma 3T and Siemens Terra 7T scanners. Both samples had equal gender distribution.

### HFGAN Construction and Method Comparison

To predict 7T images from 3T data, we tested two deep learning methods, 3D CNN [22] and 3D GAN [24]. Traditional methods only predict 2D slices, which do not provide image continuity and capture fine structure in less detail. While 3D CNNs capture structural boundaries precisely, they require well-aligned data and are less effective at enhancing contrast. 3D GANs improve contrast but do not predict structural boundaries well. Our HFGAN overcomes the limitations of either method by integrating in the generation process detailed structure learning with contrast enhancement.

First, we trained a GAN to mimic the high-contrast quality of 7T MRI images. The GAN’s generators create images similar to 7T MRIs, while the discriminators differentiate between these and ground truth 7T images. To optimize detail and contrast of the predictions, we focused on extracting and incorporating high-frequency details from the images. The HFGAN framework combines a GAN-based prediction module with a reconstruction network for enhancing image details. It extracts high-frequency information from the image data, which are then fed into the reconstruction network to learn and enhance the fine features of P7T images. The process is interactive, with each layer of the network contributing to the gradual improvement of image quality in the generated P7T images. (Suppl. Figure 1).

For whole brain assessment, we used visual and quantitative measures to evaluate if P7T images have better contrast and detail than 3T images, and how closely they match ground truth 7T imaging. We assessed whole-brain visuals, grayscale histograms, and metrics like Contrast-to-Noise Ratio (CNR), Signal-to-Noise Ratio (SNR), Peak Signal-to-Noise Ratio (PSNR), and Structural Similarity Index (SSIM). Details of these measures are provided in the Quantitative Indicators section.

### HFGAN Training Data and Image Pre-Processing

Our prediction model was trained based on the Magdeburg data consisting of 30 adult individuals. For each individual, we have both 3T T1 MPRAGE and 7T T1 MPRAGE. All 3T T1 were acquired on a Siemens Verio, with 0.8mm isotropic resolution, TR = 2500ms, and TE = 3.47ms. 7T T1 were acquired on a Siemens Magnetom 7T scanner, with 0.6mm isotropic resolution, TR = 2500ms, and TE = 2.8ms. Then we evaluated our model on an external dataset: Shanghai dataset, consisting of 43 paired 3T T1 MP2RAGE and 7T T1 MP2RAGE. For Shanghai dataset, the 3T were acquired on a Siemens Prisma scanner, with 1 mm isotropic resolution, TR =2000ms, and TE = 2.44ms. The 7T were scanned by a Siemens Terra scanner, with 0.65mm isotropic resolution, TR =3800ms, and TE = 2.36ms (Suppl. Table S2). The demographic of two datasets was listed in Extended. Table 1.

In the data preprocessing for model training, we first performed skull-stripping on MRI scans using Freesurfer [52], and then linearly aligned each 3T T1 scan to its corresponding MRI scan in the Montreal Neurological Institute (MNI) space [53]. It means 3T and 7T images have same spatial resolution, which is to ensure uniformity for model training and prediction. In consideration of field inhomogeneity, we conducted N4 correction on 7T MRI [54]. After that, the intensity values of 3T and 7T images was normalized. The final P7T images generated by the model were at the same resolution as the aligned 3T and 7T images.

Voxel-Based Morphometry (VBM) analysis was conducted using the CAT12 [36] toolbox within SPM12 to investigate structural brain differences among 3T, P7T and 7T subjects. As part of the preprocessing pipeline, T1-weighted images underwent reorientation, segmentation into grey matter (GM), white matter (WM), and cerebrospinal fluid (CSF), and normalization to the MNI space using the DARTEL (Diffeomorphic Anatomical Registration Through Exponentiated Lie Algebra) method. To enhance statistical robustness, an 8 mm full-width at half-maximum (FWHM) smoothing kernel was applied before conducting statistical analyses to examine GM volume differences.

To account for anatomical differences between populations, affine regularization was applied using different brain templates: the European brains template for the Magdeburg dataset and the East Asian brains template for the Shanghai dataset. This step ensured that morphological variations between populations were appropriately considered during normalization.

### The HFGAN network

In suppl. Figure 1, the above network called GAN network contains two components: Generator (generating P7T images) and Discriminator (discriminating P7T images with real 7T). For the generator, there are six encoders and five decoders, each of which has two 3D convolution layers [55]. Instance normalization (IN) [56], Rectified Linear Unit (Relu) [57] and Max pooling [58] were utilized (bule arrows) after each encoding processer. Meanwhile, IN, Relu and Up-convolution (Upconv) were employed (green arrows) after each decoding processer. Additionally, the numbers of encoders and decoders represent the channels of each convolution layer.

The HF information of the k-space [59] resides outside the central areas can provide anatomical details. In our implementation, two matrixes were designed and used to sample images in k-space domain (suppl. Figure 2b). Given a source image, we converted it to k-space using Fourier Transformation and then extracted HF information (suppl. Figure 2a). It is noticeable that HF images have finer structural information after repeated sampling. As shown in suppl. Figure 1, the Recon-net, aiming to learn 7T 2^nd^ HF information, has the same architecture with the generator. However, the input of the Recon-net was produced by combining the 3T 1^st^ HF and 3T 2^nd^ HF images through a convolution layer, whereas the generator takes only a 3T image. For each encoder pair of Recon-net and generator, a fusion scheme to enhance features of encoders using HF information was employed:

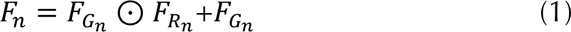

Where ⊙ indicates element-wise multiplication. *F_n_* is the fused feature of *n*-th encoder. *F_Gn_* and *F_Rn_* represent features of generator and reconstruction network respectively. For decoders, we attempted to share features of Recon-net with generator via concatenation.

### Implementation details and loss function of HFGAN

This study employed a five-fold cross-validation method with stratified sampling for data segmentation to ensure comprehensive test results from all subjects. The training, validating and testing of HFGAN were conducted on an 80GB NVIDIA A100 30C GPU with CUDA version 12.0, using Python 3.10.4 and PyTorch 1.11.0 frameworks. The network and its components were trained using the Adaptive Moment Estimation (Adam) optimizer [60] to ensure effective parameter updates and optimization. The initial learning rate was set at 0.0002, with a specific adjustment strategy where the rate was modified every 100 iterations based on a lambda rule, aimed at gradually reducing the learning rate to refine network weight adjustments. The entire training process spanned 200 epochs, combining initial rapid iteration with subsequent learning rate decay to balance early progress and later fine-tuning.

HFGAN employs a GAN network to master the image contrast characteristic of 7T images. It also incorporates a reconstruction component (Recon) specifically designed to capture the high-frequency details of 7T imaging, thereby enhancing the finer details of P7T images. For the reconstruction (R) network, We use mean square error (MSE) [61]:

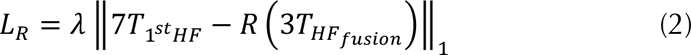

The reconstruction network within HFGAN is designed to assimilate and refine a composite of high-frequency features from 3T MRI data, denoted as *HF_fusion_*. This process involves the harmonization of both first-order high-frequency (1^st^ HF) and second-order high-frequency (2^nd^ HF) information, aligning closely with the consistency of the 1^st^ HF information derived from 7T images. Within this context, the weight coefficient *λ* of the L1 loss is crucial for maintaining balance in the loss function, and it has been calibrated to a value of 10 to optimize the learning of these intricate high-frequency details.

The generator (G) in the network optimizes its performance through a composite loss function that fuses both adversarial [62] and L1 losses:

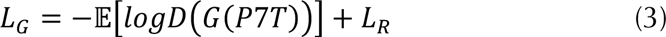

This integrated loss framework strategically combines the adversarial dynamics, compelling G to generate images that can convincingly pass for real, with the precision of L1 loss, which meticulously reduces the pixel-level differences between the generated images and their true counterparts. This synergy of loss components ensures that G not only fools the discriminator but also excels in producing high-quality reconstructions with remarkable accuracy and detail fidelity.

The discriminator within the GAN framework employs a loss function meticulously crafted to assess the authenticity of images. This function serves as a critical arbitrator, distinguishing between the high-resolution images crafted by the generator and actual 7T images.

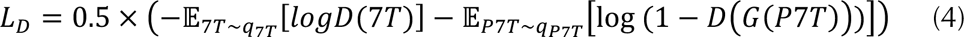

The discriminator’s loss function in the GAN architecture hinges on two critical terms. The first term is the natural logarithm of the discriminator’s output for real 7T images, drawn from the distribution *q_7T_*. This term is a measure of the discriminator’s performance in recognizing authentic 7T data, with an ideal output value of 1, reinforcing its ability to assign high probability scores to real images. The second term relates to the P7T images synthesized by the generator, reflecting the distribution *q*_2./_. For these generated images, the discriminator’s ideal output is 0, indicating its success in identifying and assigning lower probability scores to synthetic data. This bifurcated approach is designed to sharpen the discriminator’s acumen in distinguishing genuine data from fabricated equivalents.

The comprehensive loss function of HFGAN is defined as:

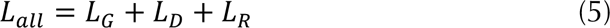

This fusion of adversarial, discriminative, and reconstruction losses orchestrates a balanced training regime. It adeptly ensures that the generator crafts images of such convincing realism that the discriminator struggles to flag them as inauthentic, while simultaneously reinforcing the structural congruence of the synthesized images with their real-life counterparts. This harmonized approach to loss not only propels the generation of visually indistinguishable images but also preserves the fidelity of the intrinsic detail characteristic of the original images.

### Quantitative Indicators

We utilized various quantitative metrics to assess both the entire brain and specific brain regions. For the entire brain analysis, we employed widely used metrics such as Peak Signal-to-Noise Ratio (PSNR) and Structural Similarity Index (SSIM) to evaluate the similarity of 3T and P7T based on the 7T groundtruth.

PSNR [63] is typically employed to compare differences between the original image and a compressed or processed version. PSNR quantifies the fidelity of an image by measuring the ratio between the maximum possible power of a signal and the power of distortion or noise present in the image. The mathematical definition of PSNR is as follows:

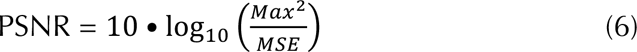

Where Max is the maximum possible value of contrast in the 3T/P7T image. MSE stands for Mean Squared Error, measuring the average squared differences between the 7T groundtruth and processed 3T/P7T image voxels. The higher the PSNR, the better the image fidelity and the closer it is to groundtruth.

SSIM [64] is a widely used metric for measuring the similarity between two images, which takes into account the luminance, contrast, and structure of the images. The mathematical definition of SSIM is as follows, considering two images x and y:

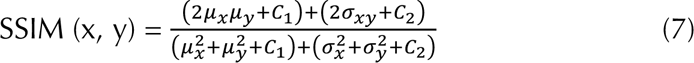

Where *µ_x_* and *µ_y_* are the means of the images x and y, respectively. 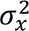 and 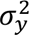 are the variance of x and y, respectively. *σ_xy_* is the covariance of x and y. *C*_1_ and *C*_2_are constants to stabilize the division with weak denominator. SSIM values range from −1 to 1, where 1 indicates the highest similarity (perfect similarity) between the images, and 0 indicates no similarity. Higher SSIM values signify a greater similarity in structural information and texture details between the compared images, implying better image quality.

For the analysis of specific brain regions, we employed three evaluation metrics: Contrast-to-Noise Ratio (CNR), Signal-to-Noise Ratio (SNR), and Dice coefficient. CNR and SNR were utilized to assess the extent to which the model enhances the contrast in the 7T predictions. On the other hand, Dice coefficient was used to evaluate the consistency between the tissue segmentations of 3T and P7T images after contrast enhancement, compared to the 7T groundtruth.

The mathematical definition of CNR [65] in the context of medical imaging is as follows:

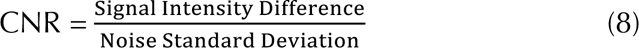

Where Signal Intensity Difference represents the difference in signal intensity between the region of interest and the surrounding background or noise. Noise Standard Deviation is the standard deviation of the noise in the image, often measured in a region with no significant signal. A higher CNR indicates a better distinction between the signal and the noise, implying better image quality for identifying the desired features or structures.

The mathematical definition of SNR [65] is typically expressed as:

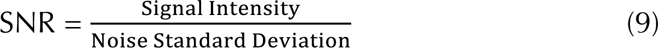

Where Signal Intensity represents the magnitude or strength of the desired signal within the region of interest in the image. Noise Standard Deviation denotes the standard deviation of the noise. A higher SNR indicates a better quality of the signal relative to the noise, implying improved image quality and better ability to discern relevant features or structures within the image.

Mathematically, if A and B are two sets representing the segmented regions in two different images (e.g., predicted and ground truth segmentations), then the Dice coefficient [66] is calculated as:

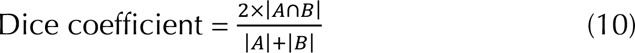

Where |*A*| represents the size or cardinality of set A. |*B*| represents the size or cardinality of set B. |*A* ∩ *B*| represents the size of the intersection of sets A and B. The Dice coefficient ranges from 0 to 1, where 0 indicates no overlap (no similarity), and 1 indicates a perfect match or complete overlap between the two sets. In the context of image segmentation, a higher Dice coefficient signifies a better agreement between the predicted segmentation and the ground truth, implying more accurate segmentation results.

### Brain measurements for case-control comparisons

To investigate case-control comparisons in the clinical cohort, we relied on widely adopted metrics like cortical thickness and subcortical volume. Recognizing HFGAN’s effect on image contrast, we also included gray-white matter contrast (GWR) to better capture variations in regional brain features.

To evaluate GWR across both cortical and subcortical regions, we extracted gray and white matter signal intensities from T1-weighted MRI images. For cortical regions, gray matter intensity was sampled at 35% depth from the gray-white matter boundary to avoid contamination by cerebrospinal fluid (CSF), ensuring anatomically consistent gray matter signals across varying cortical thicknesses. White matter intensity in cortical areas was sampled 2mm below the gray-white boundary to capture a pure white matter signal with minimal interference from adjacent gray matter [67].

In subcortical regions, gray matter intensity was sampled using region-specific masks generated from the aseg.mgz file of Freesurfer, with each structure assigned a unique label. These binary masks were applied to the T1 image to isolate the gray matter signal for each subcortical structure. Similarly, white matter intensity extraction in subcortical regions involved the creation of region-specific white matter masks, allowing precise white matter signal isolation for each structure.

The GWR for each region was calculated using the formula [68]:

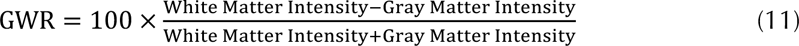

This calculation transformed gray-white matter signal differences into a percentage-based metric, with higher GWR values indicating stronger gray-white contrast and values near zero or negative suggesting reduced contrast, potentially reflecting tissue alterations. To enhance the reliability of GWR measurements, a 30 mm Gaussian smoothing filter with full-width at half-maximum (FWHM) was applied [69]. This smoothing method limited the smoothing to neighboring cortical points along the cortical mantle, thereby preserving anatomical accuracy by avoiding data blending across sulcal spaces and cerebrospinal fluid areas. This unified approach facilitated a comprehensive analysis of gray-white matter contrast, revealing potential biomarkers of psychiatric and neurological changes across different brain structures.

### Statistical analysis

In this study, paired *t*-tests [70] were used for all group comparisons base on SPSS Statistics 25 and SPM12. Specifically, in Figure 3, *t*-tests were performed to assess differences across brain regions in the datasets. In Suppl. Figure 4, *t*-tests were employed to compare gray matter volumes (adjusted for intracranial volume) among 3T, P7T and 7T. Likewise, in Suppl. Figures 6, 7, 8, and 9, *t*-tests were used to evaluate differences in Dice coefficients, subcortical/cortical measures, and other metrics. Unless otherwise stated, statistical significance was defined at P < 0.05.

For all case-control comparisons, we employed a generalized linear model (GLM) [71] by MATLAB R2020b. In this model, the measurement from each brain region (e.g., cortical thickness or gray–white matter ratio) served as the dependent variable (Y), while group membership (e.g., case vs. control) was the independent variable (X). Additional covariates (e.g., age, gender, scanning site, weight) were included to mitigate potentially confounding effects, ensuring that observed differences in the dependent variable primarily reflected the influence of group status. All P-values were corrected for multiple comparisons within each modality using the Benjamini–Hochberg false discovery rate (BH-FDR) procedure [72], and statistical significance was defined at P< 0.05 unless otherwise stated.

To calculate the proportion of explained variance (partial η^2^), we employed a General Linear Model (GLM) to examine group differences in aggregated brain measurements (e.g., weighted averages of cortical thickness or gray–white matter ratios), while controlling for relevant covariates (e.g., age, gender, and scanning site). To quantify the unique contribution of the group variable after adjusting for these covariates, we computed the partial η^2^ using the following formula:

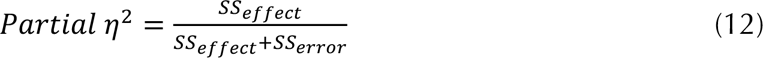

Where *SS_effect_* is the sum of squares for the effect of the group variable, and *SS_error_* is the sum of squares for the residual error. All P-values were adjusted for multiple comparisons via BH-FDR, with P < 0.05 considered statistically significant.

### STRATIFY/ESTRA

The STRATIFY and ESTRA studies enlisted individuals between the ages of 19 and 25 who were diagnosed with alcohol use disorder or major depression disorder (STRATIFY), anorexia nervosa or bulimia nervosa (ESTRA), as well as a control group without any mental disorder diagnoses. Recruitment took place at three locations: Berlin, London (London has two sites: CNS and Invicro), and Southampton. Gender distribution, based on self-identification, showed variation among the different groups with mental health disorders (see Extended. Table 3 for details). In our analysis of data from both STRATIFY and ESTRA, variables such as recruitment site and gender were accounted for as covariates. Detailed demographic information can be found in Extended. Table 3. Additionally, the scanning protocol can be seen in Suppl. Table S5

### ADNI2

ADNI2 (Alzheimer’s Disease Neuroimaging Initiative Phase II) is a multicenter study aimed at advancing the research initiated in ADNI1 through the use of more sophisticated biomarkers, imaging technologies, and data collection methods. ADNI2 introduces novel testing techniques, such as amyloid-beta and tau protein PET scans, along with enhanced MRI technology. Furthermore, it extends research to include cognitively normal elderly individuals, to explore early changes in cognitive function and gain a deeper understanding of the progression of Alzheimer’s disease. This phase of the study enriches our understanding of the biological underpinnings of Alzheimer’s and related conditions, supporting the development of new treatment approaches and early diagnostic tools. The study encompasses participants categorized as progressive mild cognitive impairment (pMCI, N=86), stable mild cognitive impairment (sMCI, N=224), Alzheimer’s disease (AD, N=147), and healthy controls without neurodegenerative diseases (HC-2, N=186). We enhanced T1 structural images to produce P7T images via HFGAN. For categorization, pMCI and sMCI are grouped as MCI, and together with AD, are classified as PAT-2. Demographic details are provided in Extended. Table 4. Additionally, the scanning protocol can be seen in Suppl. Table S5

## Supplemental Materials

For our quantitative analysis, we compared the similarity between P7T and real 7T images using PSNR and SSIM metrics (Extended. Table 2). HFGAN outperformed 3D CNN and 3D GAN, showing higher mean PSNR and SSIM values across datasets, which indicates better similarity with actual 7T images. HFGAN also had the smallest standard deviations, demonstrating its stable and consistent prediction capabilities.

### Case-control comparisons (STRATIFY/ESTRA) based on cortical GWR and sub-cortical volume

We also performed case-control comparisons of cortical GWR and subcortical volume in STRATIFY/ESTRA (Suppl. Figure 11 and Suppl. Table S6). Regarding intergroup differences in cortical GWR, P7T identified significant differences in the left ER, FF, IT, MT, Pop and right LIN between AUD patients and healthy subjects (FDR < 0.05). In contrast, 3T cortical thickness was completely insensitive to the brain region differences between AUD and HC-1.

For ED, left FF, POC, SP, TT and right SP showed significant differences with P7T (FDR < 0.05 or < 0.01), while 3T detected no significant differences (Suppl. Figure 11c and Suppl. Table S6). Consistent with the thickness results, neither 3T nor P7T brain region contrast could detect significant changes in MDD (Figure 11d and Suppl. Table S6).

When analyzing subcortical volume, neither P7T nor 3T identified any significant differences in subcortical regions within the control groups (Figure 11e and Suppl. Table S6).

### Case-control comparisons (ADNI2) based on cortex thickness and sub-cortical volume

In subcortical volume measurements (Suppl. Figure 14b), P7T found additional significant differences in bilateral CAU (Left, FDR < 0.01; Right, FDR < 0.05) and in right THA (FDR< 0.01) when comparing HC-2 to PAT-2. In HC-2 vs. MCI, P7T identified additional significant differences in bilateral THA (Left, FDR < 0.05; Right, FDR < 0.05). In comparisons between HC-2 and AD patients, P7T also found additional significant differences in bilateral CAU (Left, FDR < 0.0001; Right, FDR < 0.01). When distinguishing between MCI and AD patients, P7T found significance in bilateral CAU (Left, FDR < 0.001; Right, FDR < 0.05). Between sMCI and healthy controls, P7T detected more brain region differences, primarily in the bilateral THA (Left: FDR < 0.05; Right: FDR < 0.05). Compared to sMCI, pMCI showed additional differences only in the left CAU (FDR < 0.05). Thus, P7T offers new insights into distinguishing between sMCI and pMCI.

In cortical thickness comparisons between healthy controls and more advanced disease groups such as Patients, AD, and pMCI, both 3T and P7T identified a comparable number of brain regions with significant differences. However, during the early diagnostic stages, such as in MCI, P7T (22/34) detected differences in more left cortical regions compared to 3T (18/34). Similarly, in sMCI, P7T identified more significant cortical differences than 3T in both the left cortex (10/34 vs. 8/34) and the right cortex (8/34 vs. 5/34). Notably, in early diagnostic stages, P7T revealed new significant differences in brain regions such as the lateral occipital cortex, lingual gyrus, parstriangularis, and insula as shown in Suppl. Figure14(c).

---

**Extended Table 1:** The demographics of Magdeburg and Shanghai datasets.

| Dataset | Magdeburg, N=30 |  | Shanghai, N=43 |  |
| --- | --- | --- | --- | --- |
| T1 Modality | MPRAGE |  | MP2RAGE |  |
| demographic |  |  |  |  |
|  | Mean | SD | Mean | SD |
| Age | 25.41 | 4.97 | 26.00 | 3.12 |
|  | N | % | N | % |
| Sex(Female) | 15 | 50 | 22 | 51.16 |
| 3T-7T match or not | Yes |  | Yes |  |

**Extended Table 2:** The PSNR and SSIM (Mean ± Standard deviation) of the whole brain for Magdeburg and Shanghai datasets.

| Method | PSNR |  | SSIM |  |
| --- | --- | --- | --- | --- |
|  | Magdeburg<br>(30 cases) | Shanghai<br>(43 cases) | Magdeburg<br>(30 cases) | Shanghai<br>(43 cases) |
| 3T | 21.84±3.97 | 23.78±3.18 | 21.84±3.97 | 23.78±3.18 |
| 3D CNN[22] | 23.92±3.10 | 24.85±3.00 | 19.12±3.69 | 19.99±3.04 |
| 3D GAN[24] | 26.10±2.81 | 28.90±2.69 | 20.54±2.61 | 21.20±2.63 |
| HFGAN(ours) | **30.33±1.58** | **31.22±1.18** | **23.20±0.19** | **23.65±1.21** |

Mean ↑ ± Standard deviation ↓ ;
3D CNN [22]: Widely used in medical imaging tasks;
3D GAN [24]: Widely used in image cross-modal synthesis tasks.

**Extended Table 3.** Characteristics of the STRATIFY/ESTRA cohort.

|  | Total number | F/M | Scanning sites(Num) |  |  |  |
| --- | --- | --- | --- | --- | --- | --- |
|  |  |  | Berlin | London-CNS | London-Invicro | Southampton |
| Patients-1 | 405 | 306/99 | 23 | 130 | 148 | 104 |
| AUD | 137 | 80/57 | 8 | 28 | 52 | 49 |
| MDD | 160 | 118/42 | 15 | 33 | 57 | 55 |
| ED | 108 | 108/0 | 0 | 69 | 39 | 0 |
| Controls-1 | 266 | 152/114 | 86 | 112 | 20 | 48 |

AUD, alcohol use disorder; MDD, major depression disorder; ED, Eating disorders (composed of AN and BN); AN, anorexia nervosa; BN, bulimia nervosa; PAT-1, Patients-1, combinations of multiple disorders; HC-1, Controls-1.

**Extended Table 4.** Characteristics of the ADNI2 cohort.

|  |  | Total number | F/M | Age |  |
| --- | --- | --- | --- | --- | --- |
|  |  |  |  | Mean | SD |
| Patients-2 |  | 457 | 198/259 | 72.84 | 7.71 |
| MCI | pMCI | 86 | 38/48 | 72.63 | 7.29 |
|  | sMCI | 224 | 99/125 | 71.76 | 7.39 |
| AD |  | 147 | 61/86 | 74.61 | 8.14 |
| Controls-2 |  | 186 | 96/90 | 73.52 | 6.28 |

MCI, Mild Cognitive Impairment; pMCI, Progressive Mild Cognitive Impairment; sMCI, Stable Mild Cognitive Impairment; AD, Alzheimer's Disease; PAT-2, Patients-2, combinations of multiple diseases; HC-2, Controls-2.

