## Supplementary Figure 1 for "Leveraging Deep Learning to Enhance MRI for Brain Disorders"

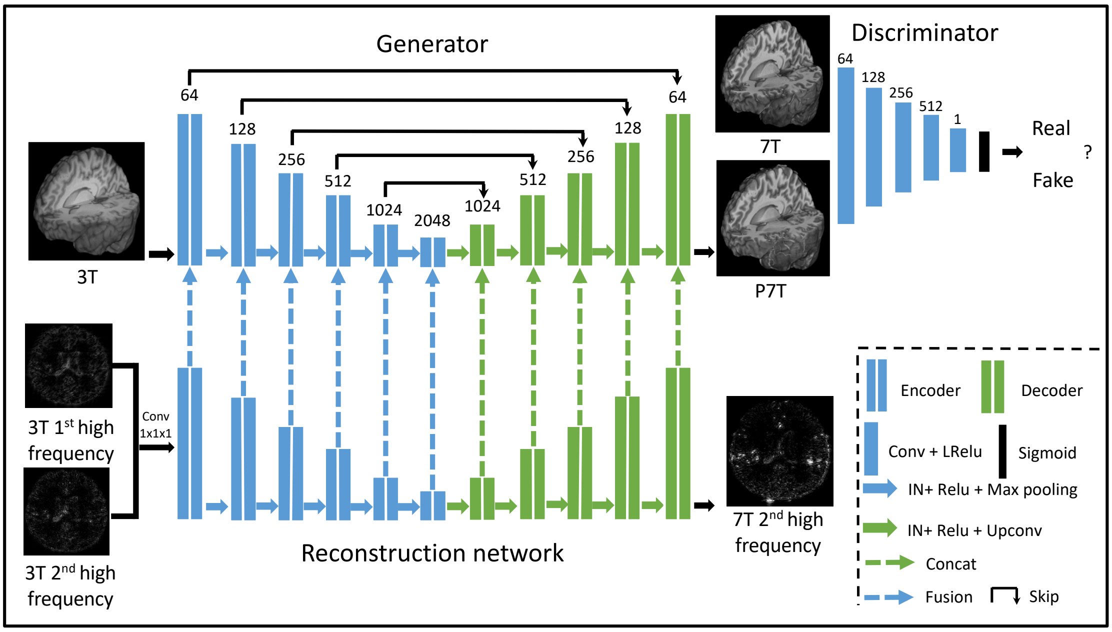


Supplemental Figure 1: The specific structure of the HFGAN. Detailed information can be seen in Suppl. Table S1.
