## Supplementary Figure 2 for "Leveraging Deep Learning to Enhance MRI for Brain Disorders"

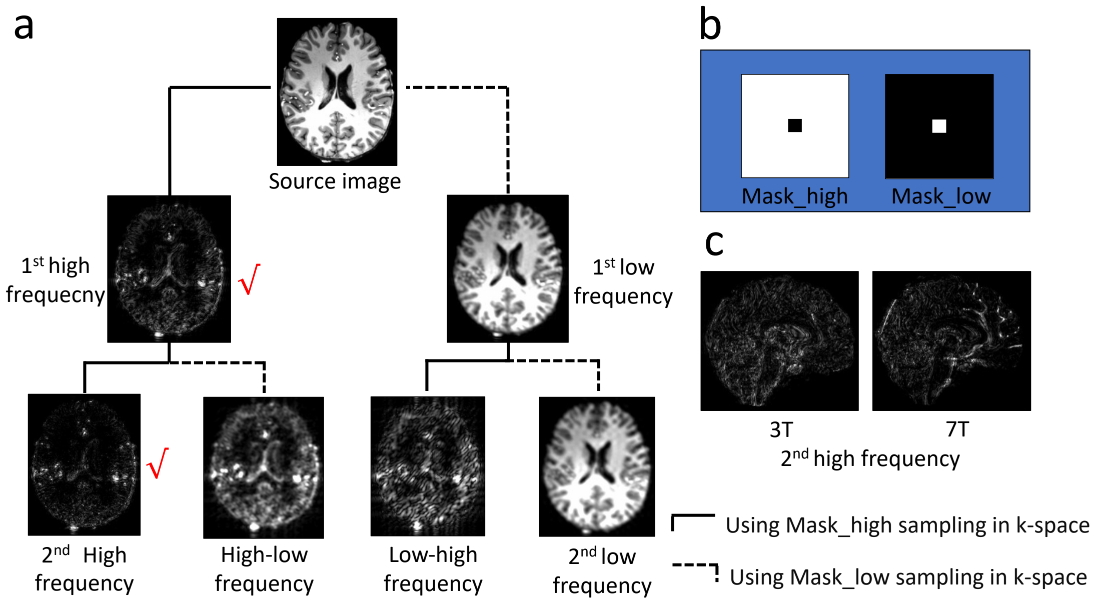


Supplemental Figure 2: a The procedure of extracting high frequency information. b The two matrices designed to sample images in k-spaces domain. C The examples of 2nd 3T and 7T high frequency, and 7T has less redundant information.
