## Supplementary Figure 3 for "Leveraging Deep Learning to Enhance MRI for Brain Disorders"

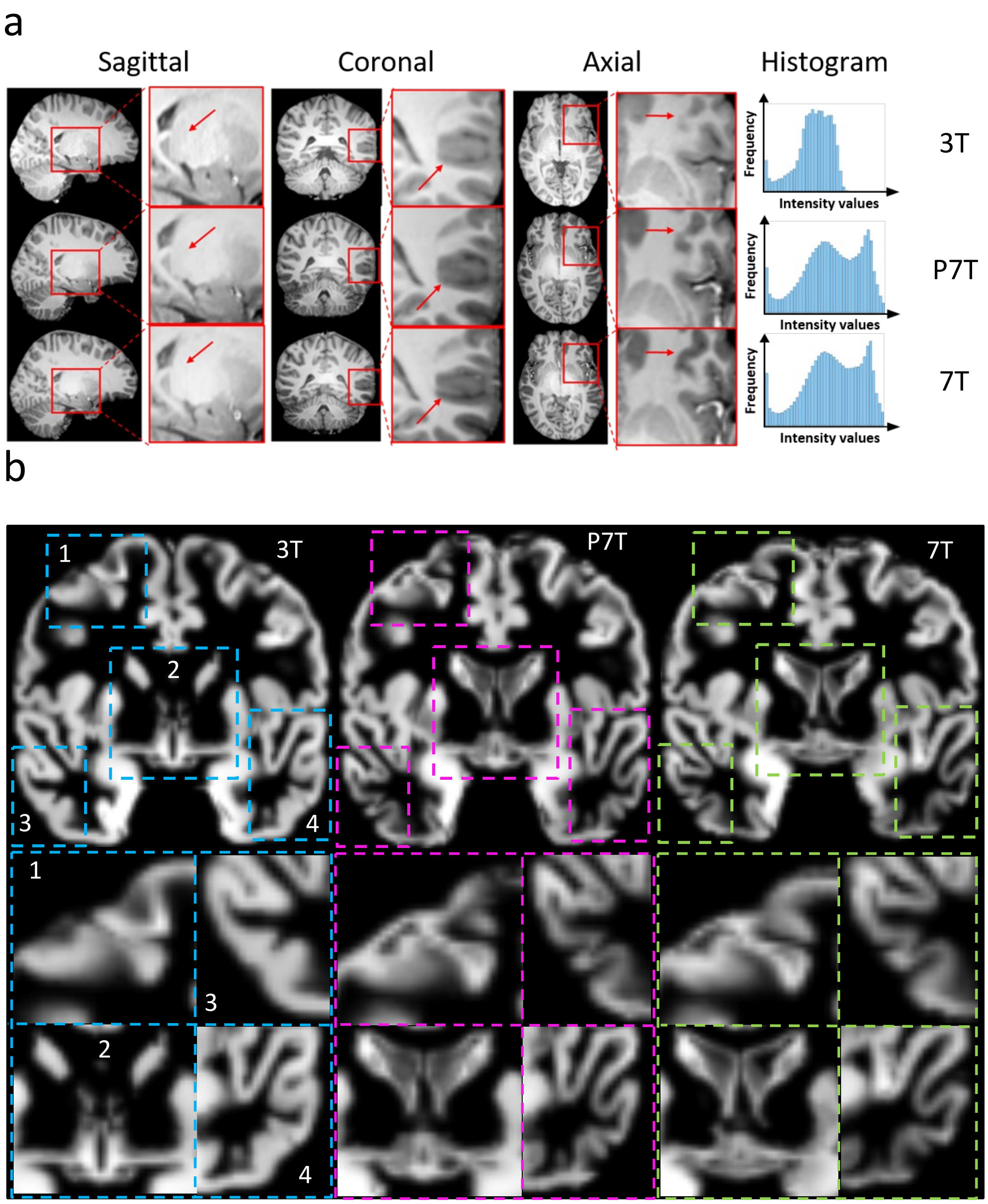


Supplemental Figure 3: a A testing case for 3T, P7T prediction and 7T of Fudan dataset which have been showcased in all three views, along with their corresponding local regions. Additionally, the grayscale histogram distribution was depicted in the accompanying figure. b The gray matter segmentation for the testing case.
