## Supplementary Figure 4 for "Leveraging Deep Learning to Enhance MRI for Brain Disorders"

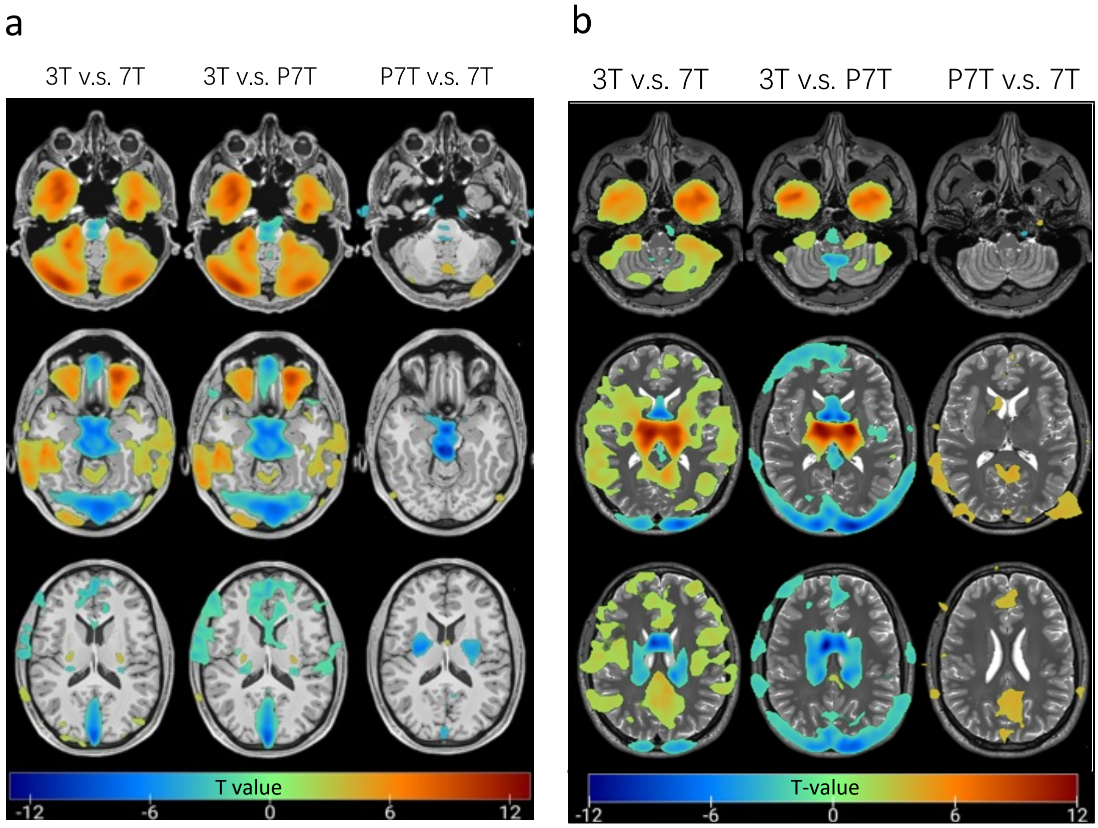


Supplemental Figure 4: We performed VBM on Magdeburg and Shanghai datasets to compare gray matter volumes from segmented whole-brain images, adjusting for intracranial volume. We used t-tests to identify differences between 3T and 7T (3T vs. 7T) and between P7T and 7T (P7T vs. 7T). The T-values showing these differences were then mapped onto the standard templates for visualization. a The Magdeburg dataset composed of T1 MPRAGE images. b The Shanghai dataset composed of T1 MP2RAGE images.
