## Supplementary Figure 5 for "Leveraging Deep Learning to Enhance MRI for Brain Disorders"

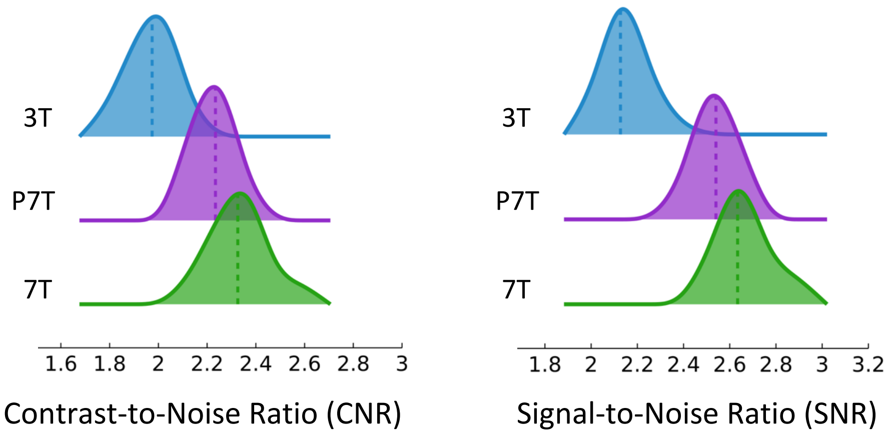


Supplemental Figure 5: The CNR and SNR assessments for segmented hippocampus of all Magdeburg cases. Here we used 7T manually segmented hippocampus.
