## Supplementary Figure 6 for "Leveraging Deep Learning to Enhance MRI for Brain Disorders"

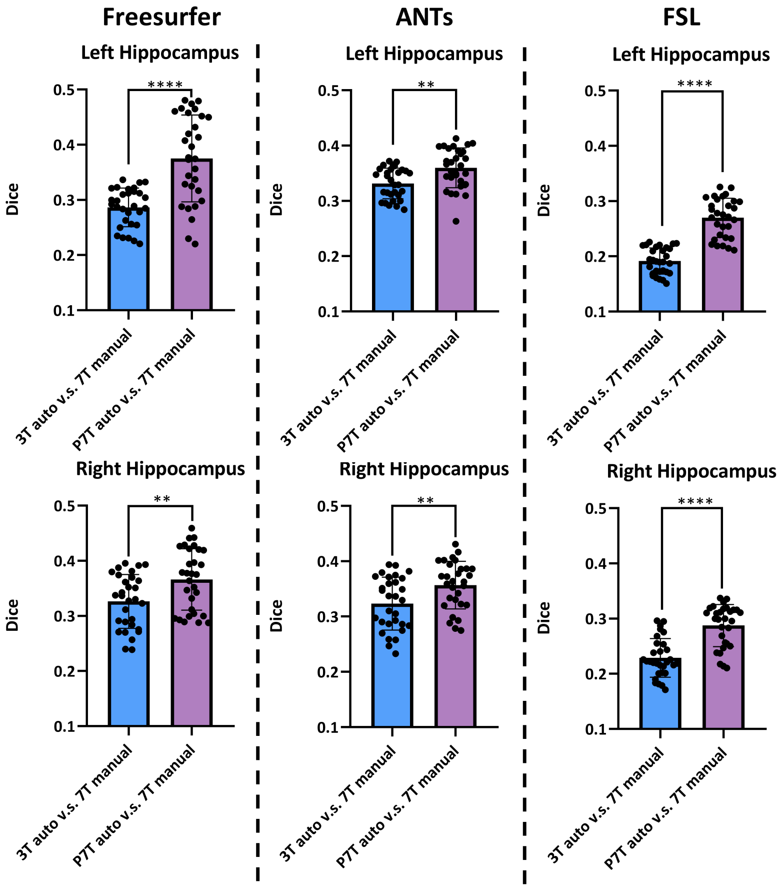


Supplemental Figure 6: Accuracy of Various Tools (Freesurfer, ANTs, FSL) for Hippocampal Segmentation. Using manually segmented hippocampus as the gold standard, we computed Dice coefficients for automated segmentation at 3T and P7T. We then employed paired t-test to differentiate the Dice distributions between 3T and P7T, enabling a comparison of the disparities between the two. **: FDR < 0.05; ****: FDR < 0.0001.
