## Supplementary Figure 7 for "Leveraging Deep Learning to Enhance MRI for Brain Disorders"

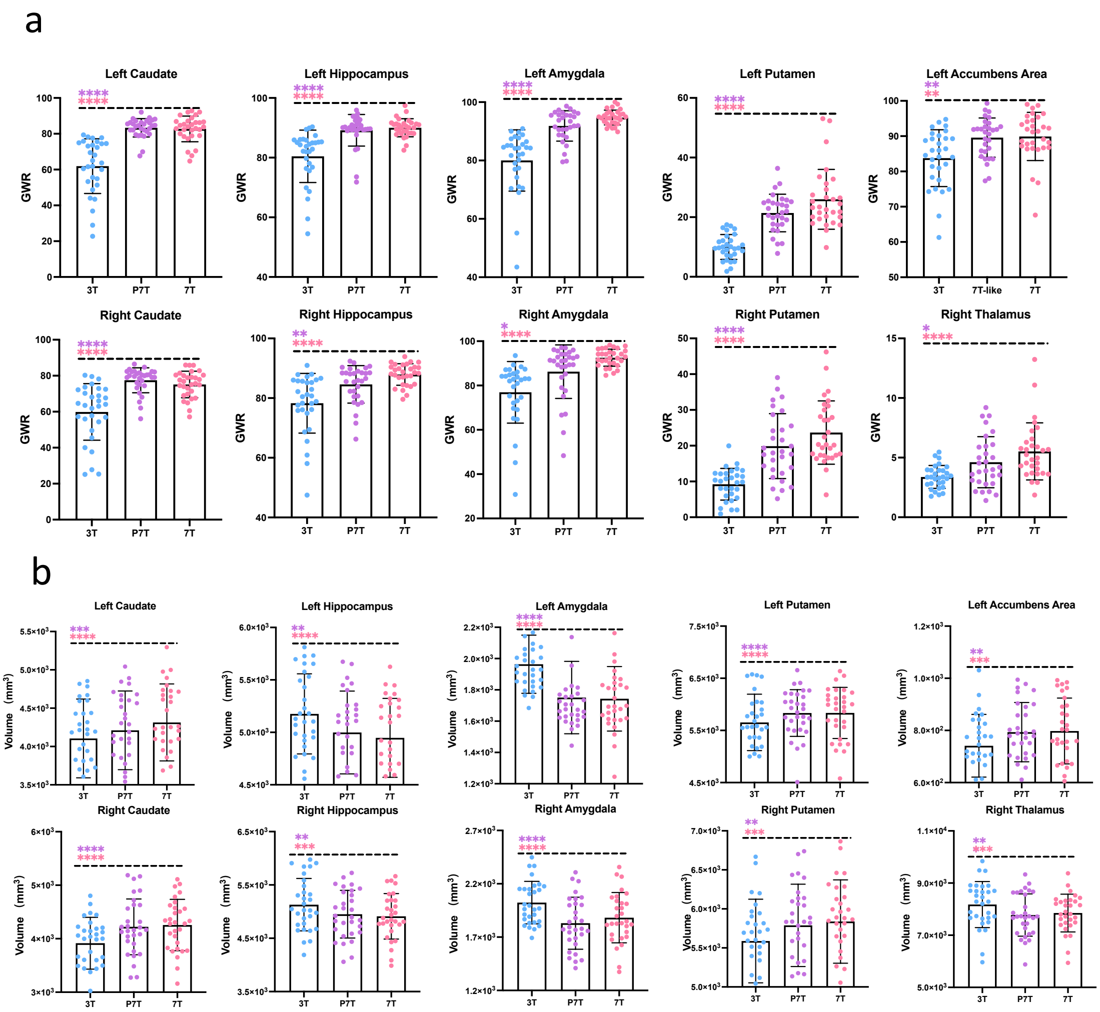


Supplemental Figure 7: Two metrics of significantly enhanced sub-cortical regions. We performed paired t-test for three groups. a The GWR difference of sub-cortical regions. b The volume difference of sub-cortical regions. Scatter plots and bar charts were simultaneously displayed, with clear annotations indicating the differences between various data and 7T MRIs. In certain brain regions, there are remarkably significant differences in voxel intensity values and volume between 3T MRIs and 7T MRIs. However, there is no significant difference between P7T predictions and 7T MRIs among these regions.
