## Supplementary Figure 8 for "Leveraging Deep Learning to Enhance MRI for Brain Disorders"

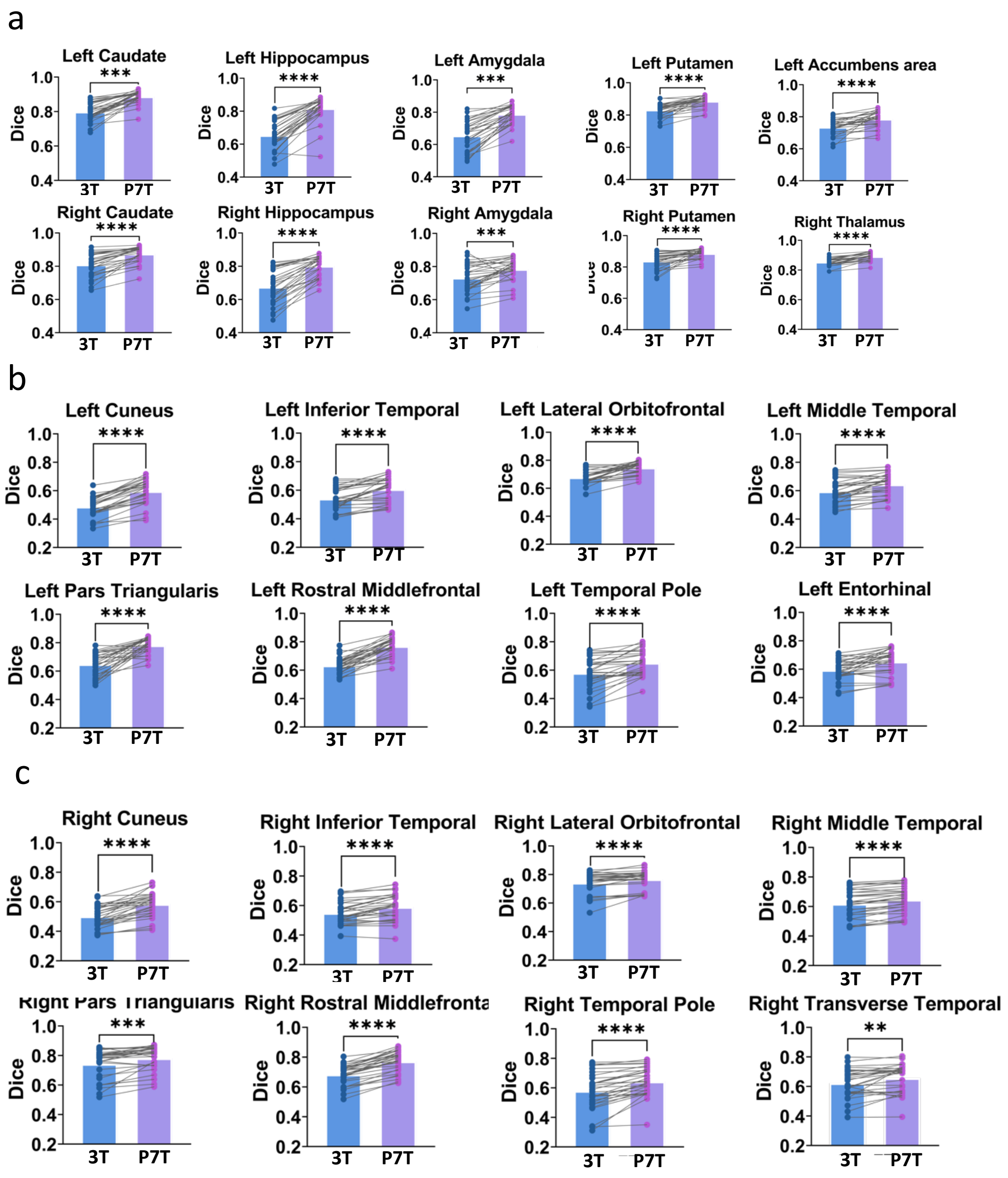


Supplemental Figure 8: We calculated the dice of 3T and P7T compared with 7T ground truth. In addition, we visualized the individual dice comparison and statistical difference between 3T and P7T. a The dice results of sub-cortical regions. b The dice results of left hemisphere. c The dice results of right hemisphere.
