## Supplementary Figure 9 for "Leveraging Deep Learning to Enhance MRI for Brain Disorders"

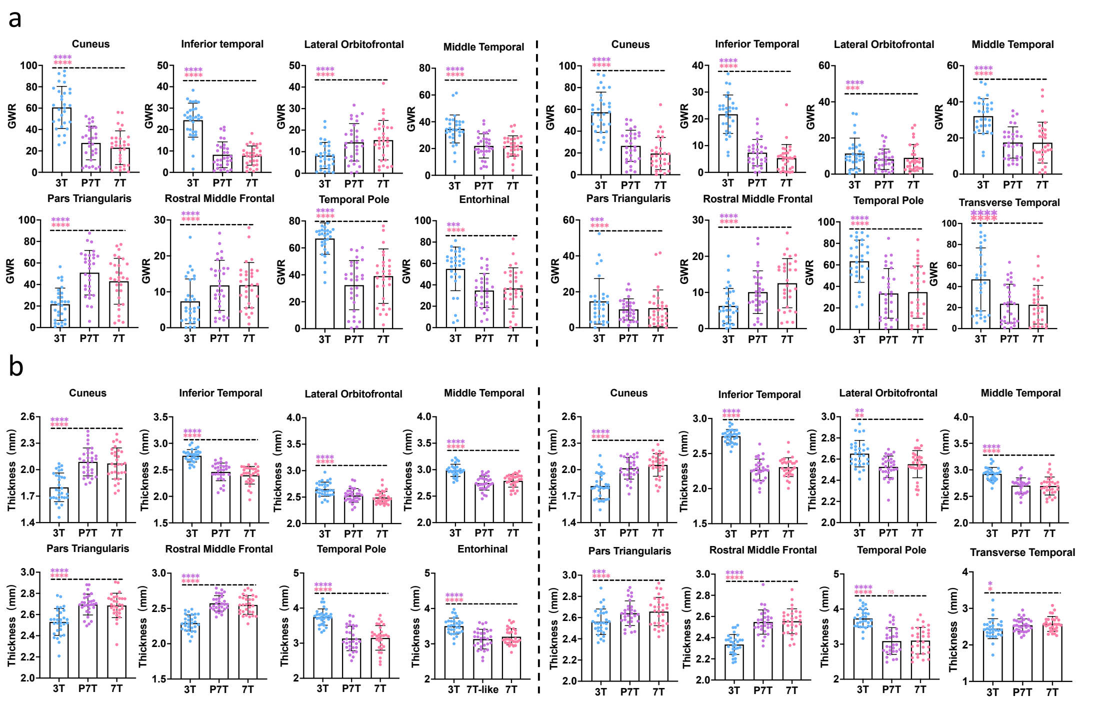


Supplemental Figure 9: Two metrics of significantly enhanced cortical regions. We performed paired t-test for three groups. a The GWR difference of left and right hemispheres. b The thickness difference of left and right hemispheres. Scatter plots and bar charts were simultaneously displayed, with clear annotations indicating the differences between various data and 7T.
