## Supplementary figures and images for "Leveraging Deep Learning to Enhance MRI for Brain Disorders"

### Supplementary Figure 10

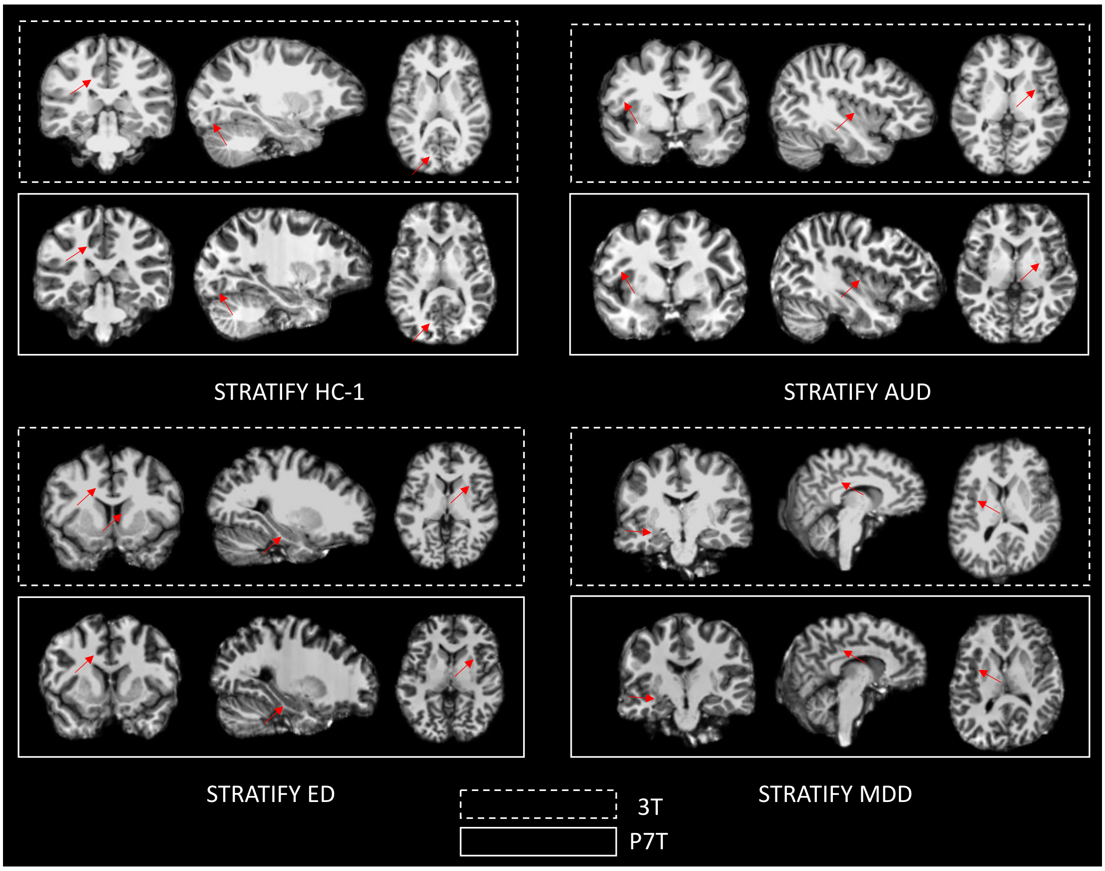

Supplemental Figure 10: The STRATIFY/ESTRA samples of 3T and P7T. We present the HC-1, AUD, ED and MDD.

### Supplementary Figure 13

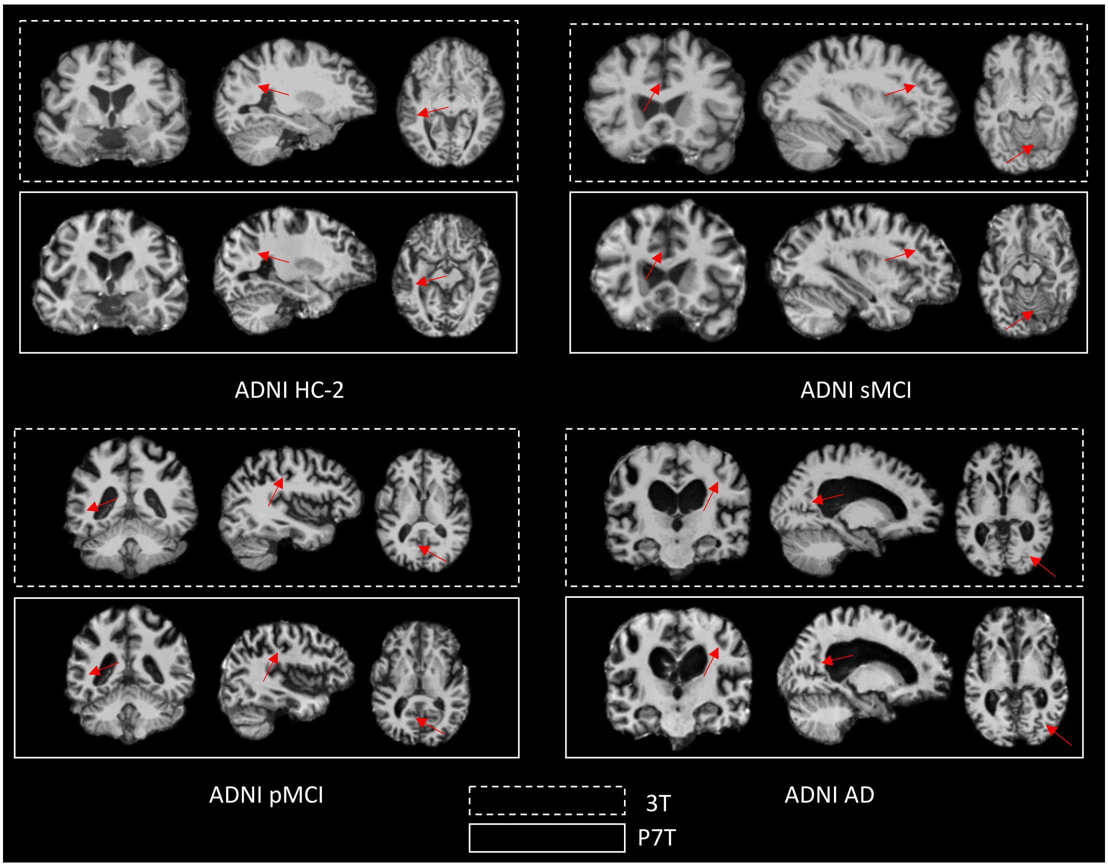


Supplemental Figure 13: The ADNI samples of 3T and P7T. We present the HC-2, sMCI, pMCI and AD.
