## Supplementary Figure 11 for "Leveraging Deep Learning to Enhance MRI for Brain Disorders"

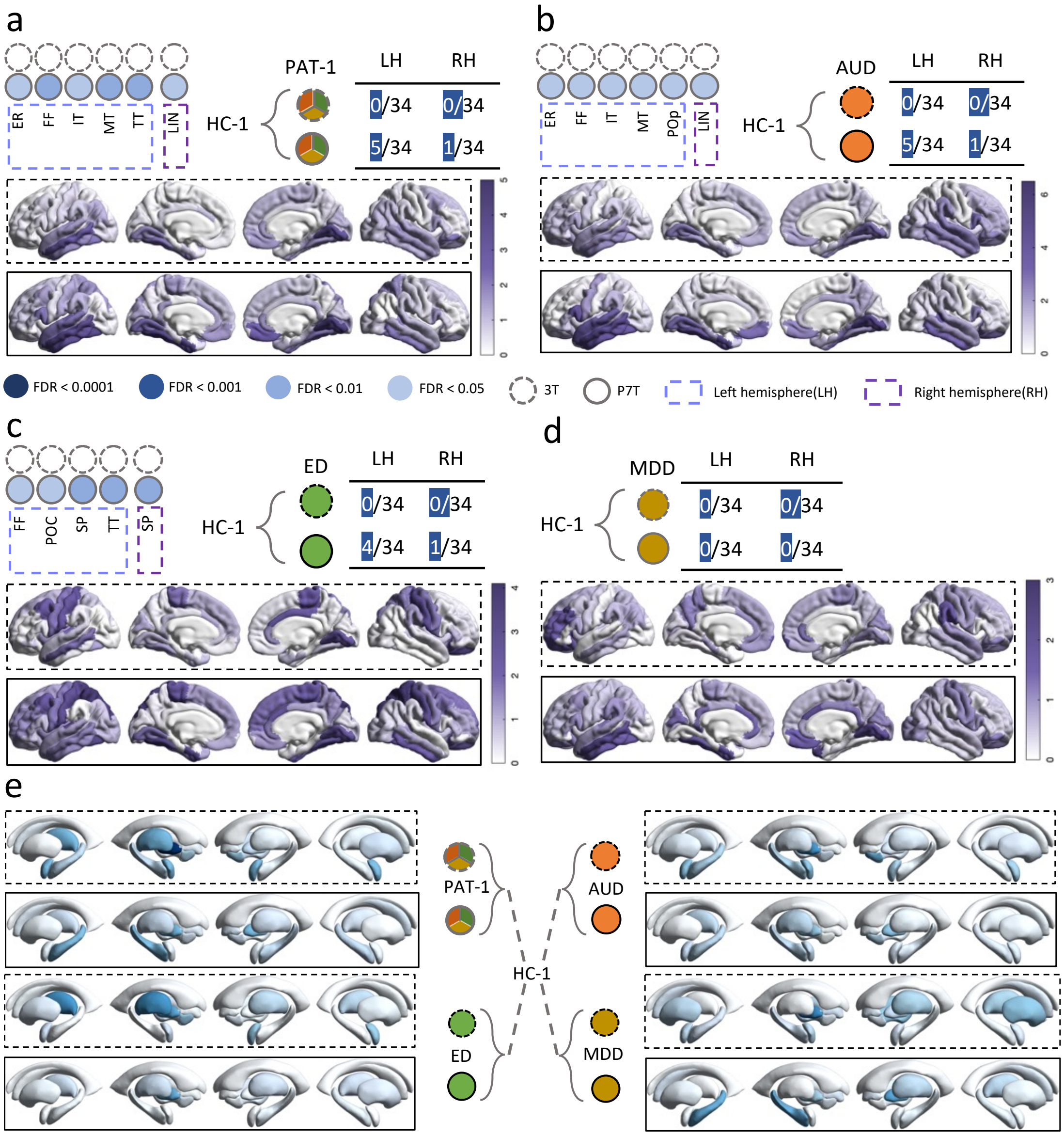


Supplemental Figure 11: The application of STRATIFY/ESTRA. We used a generalized linear model (GLM) to compare cortical gray-white matter ratio (GWR) or volume among several disorder groups and healthy controls (HC-1), adjusting for factors like age, gender, and scanning sites. For cortical GWR, we show significant FDR of regions, the proportion of significant brain regions and the absolute T-values mapped onto brain. For sub-cortical volume, we present the absolute T-values mapped onto brain There is no significant difference in sub-cortical volume. a Cortical comparison between HC-1 and PAT-1. b Cortical comparison between HC-1 and AUD. c Cortical comparison between HC-1 and ED. d Cortical comparison between HC-1 and MDD (MDD has no regions with significant difference). e Sub-cortical comparisons across all disorder groups.
