## Supplementary Figure 12 for "Leveraging Deep Learning to Enhance MRI for Brain Disorders"

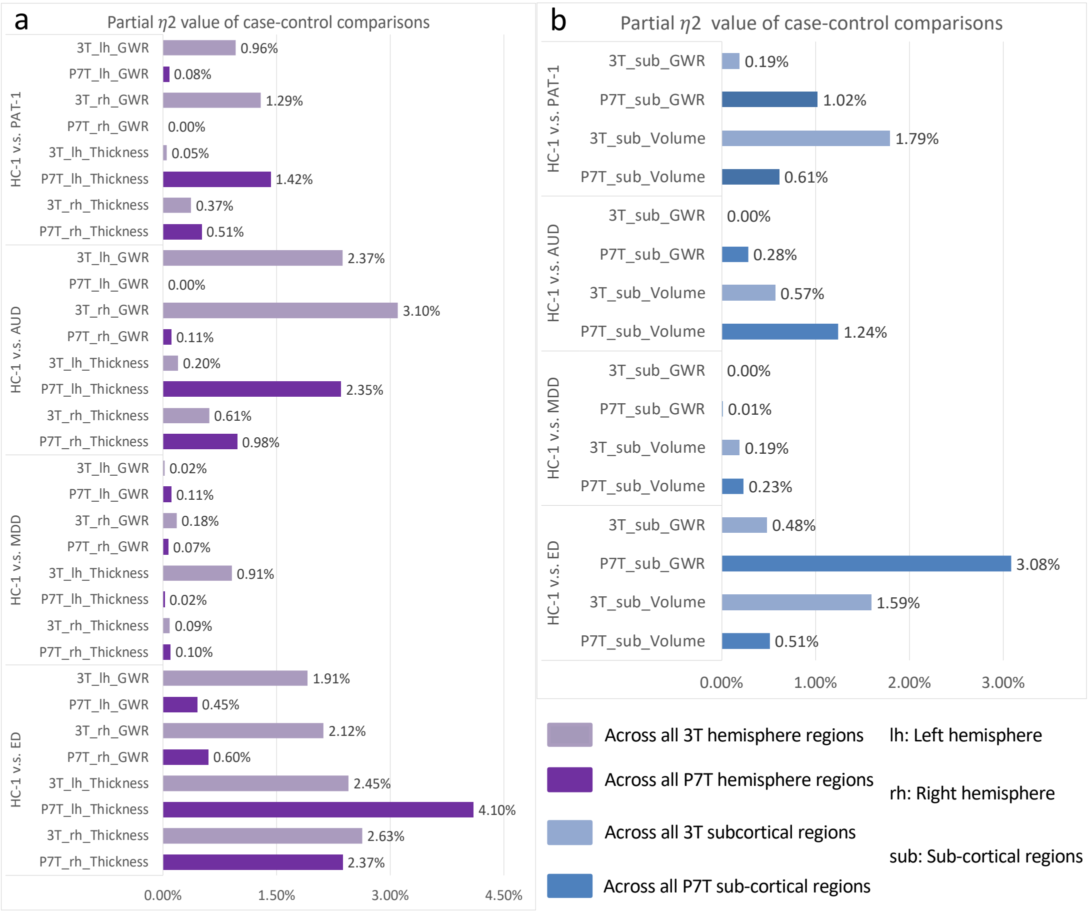


Supplemental Figure 12: In the STRATIFY/ESTRA study, we also calculated the proportion of explained variance of overall cortical and subcortical structures regarding differences between psychiatric patients and healthy controls, represented by Partial $\eta^2$ values (calculated as described in the statistical analysis section of the Methods). a Partial $\eta^2$ values represent the proportion of explained variance of different cortical measurements for disease differences. Light purple indicates 3T cortex, while dark purple represents P7T cortex. b Partial $\eta^2$ values represent the explanatory power of different subcortical measurements for disease differences. Light blue indicates 3T subcortical structure, while dark blue represents P7T subcortical regions.
