## Supplementary Figure 14 for "Leveraging Deep Learning to Enhance MRI for Brain Disorders"

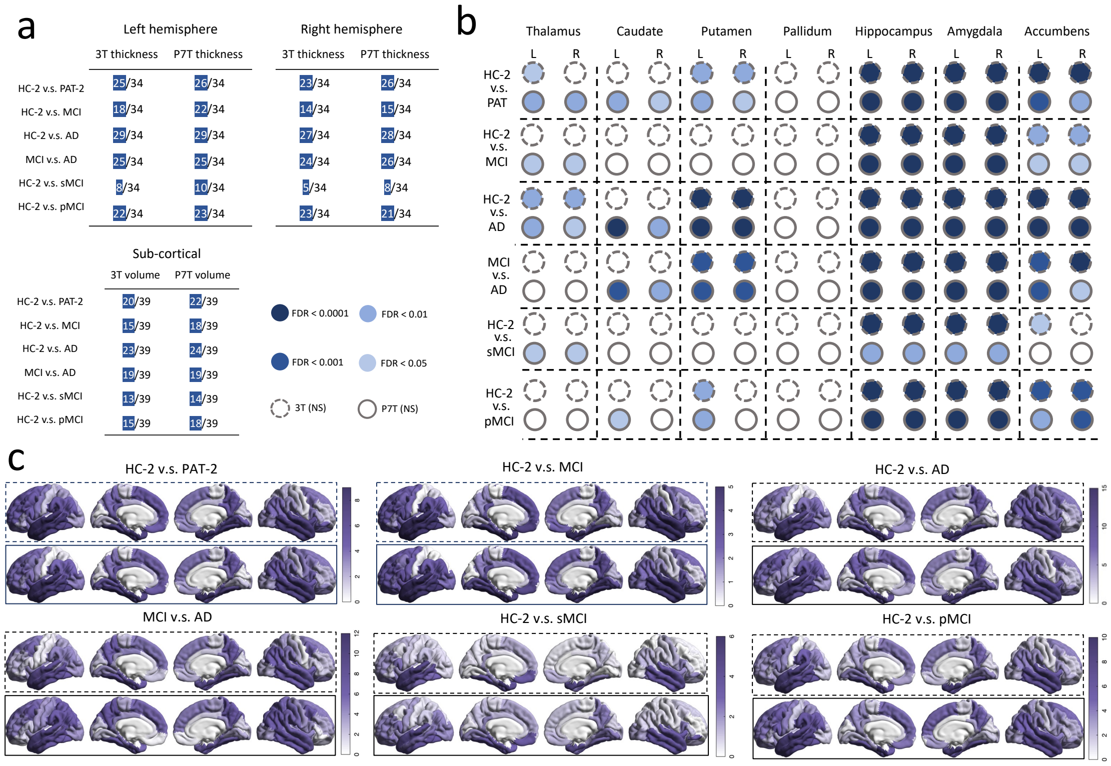


Supplemental Figure14: We examined differences between diseases and control groups in significantly affected brain areas. By measuring cortical thickness and sub-cortical volume, we used GLM to adjust for factors like age, weights and gender, enabling us to compare regional differences across the groups. a We show the number of cortical regions with significant differences for case-control comparisons. b We show significant FDR differences in sub-cortical regions of GWR. c We show the number of cortical regions with significant differences and the statistical absolute T values mapped onto cortical regions.
