## Supplementary Figure 15 for "Leveraging Deep Learning to Enhance MRI for Brain Disorders"

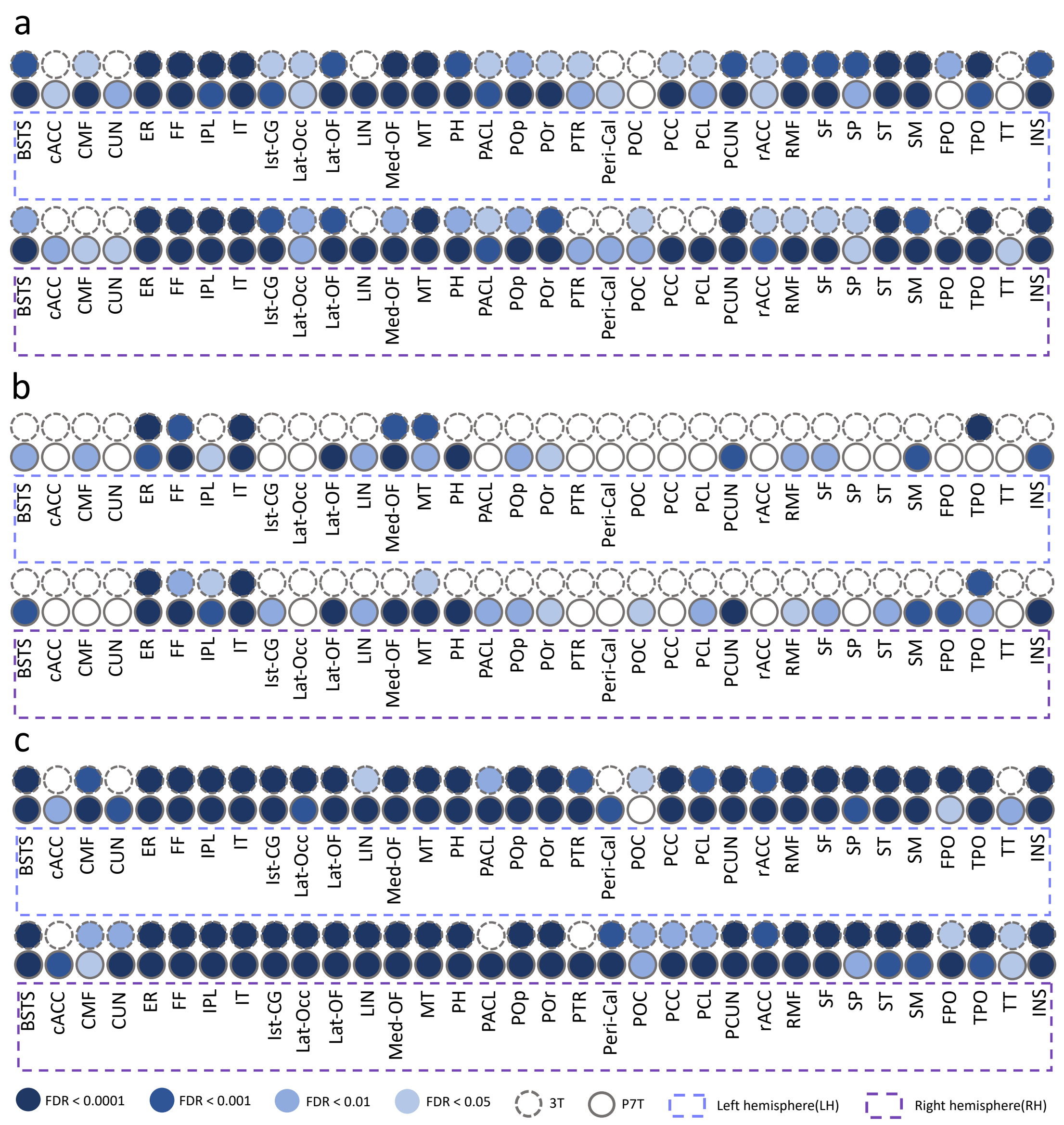


Supplemental Figure15: FDR results of ADNI GWR. a FDR between HC-2 and PAT-2. b FDR between HC-2 and MCI. c FDR between HC-2 and AD.
