## Supplementary Figure 16 for "Leveraging Deep Learning to Enhance MRI for Brain Disorders"

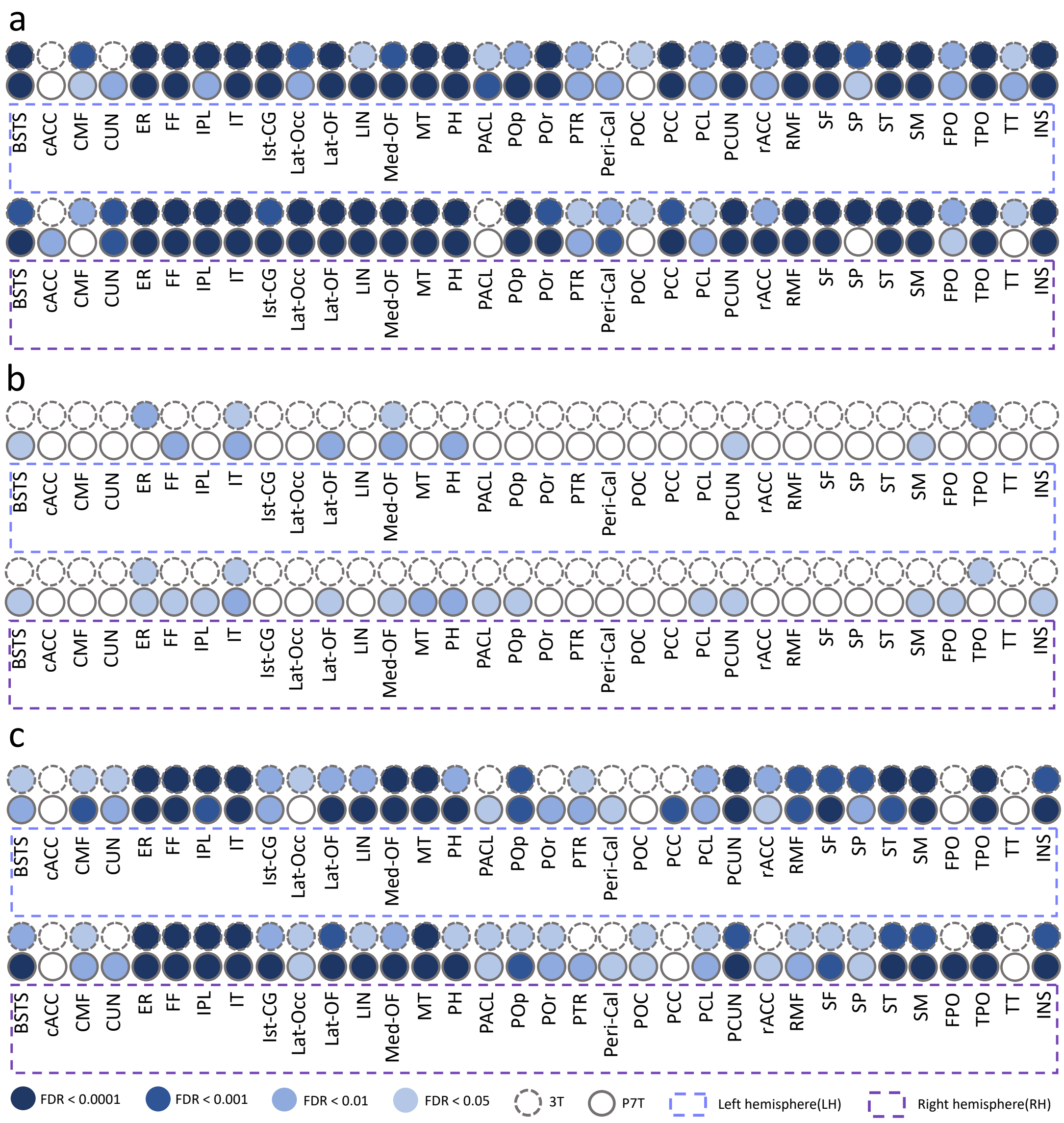


Supplemental Figure16: FDR results of ADNI GWR. a FDR between MCI and AD. b FDR between HC-2 and sMCI. c FDR between HC-2 and pMCI.
