## Supplementary Figure 17 for "Leveraging Deep Learning to Enhance MRI for Brain Disorders"

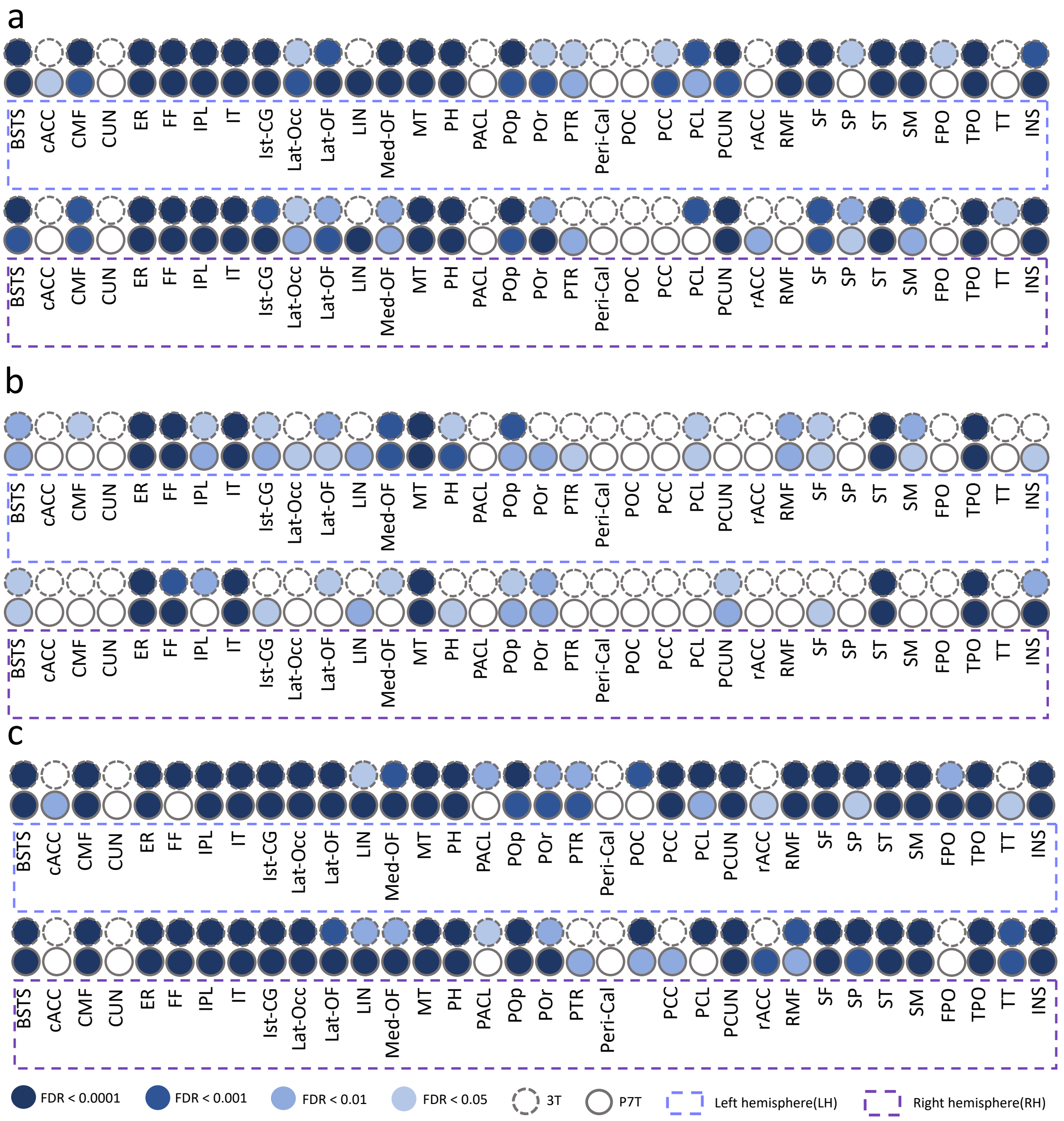


Supplemental Figure17: FDR results of ADNI thickness. a FDR between HC-2 and PAT-2. b FDR between HC-2 and MCI. c FDR between HC-2 and AD
