## Supplementary Figure 18 for "Leveraging Deep Learning to Enhance MRI for Brain Disorders"

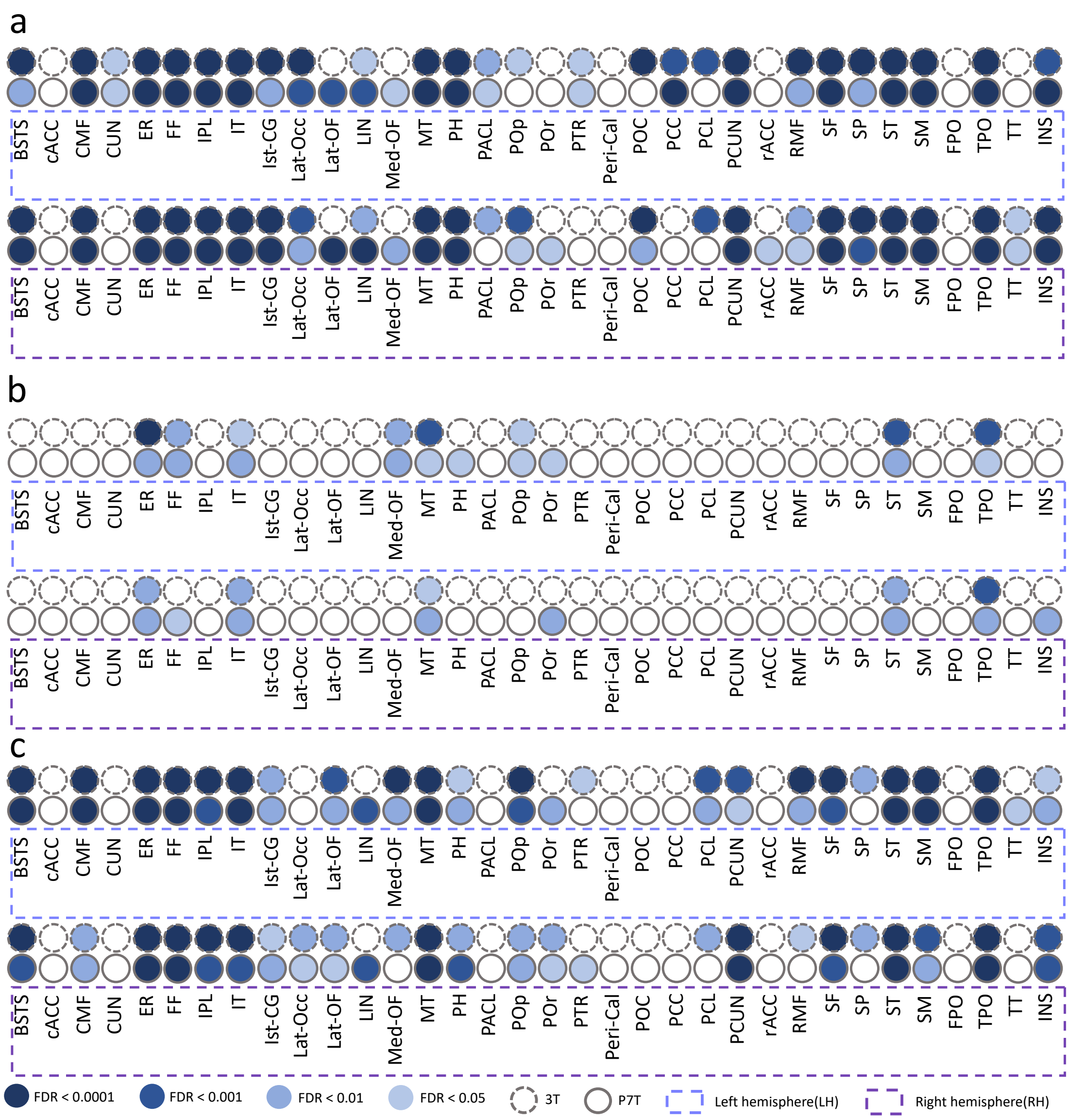


Supplemental Figure18: FDR results of ADNI thickness. a FDR between MCI and AD. b FDR between HC-2 and sMCI. c FDR between HC-2 and pMCI.
