## Supplementary Figure 19 for "Leveraging Deep Learning to Enhance MRI for Brain Disorders"

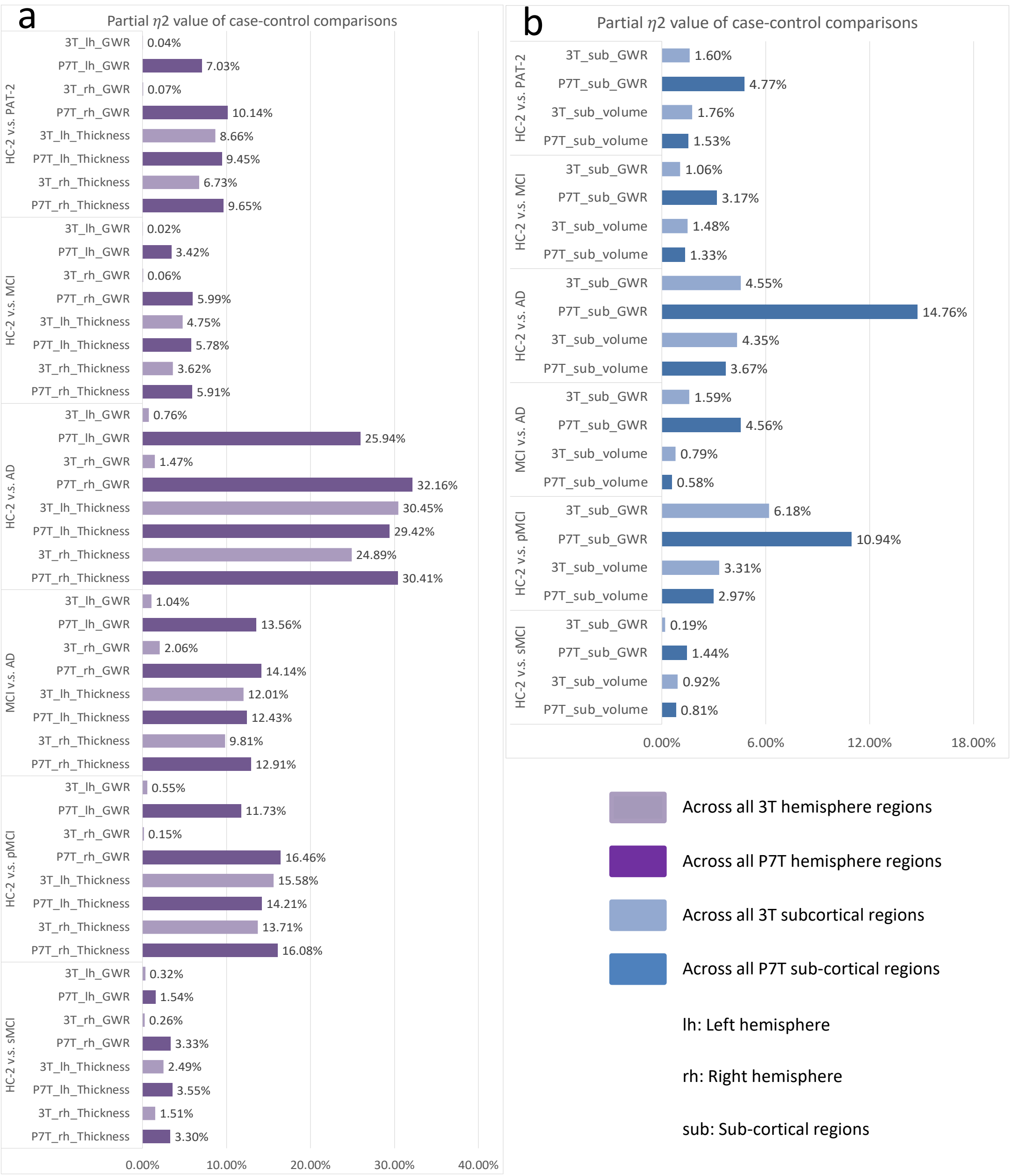


Supplemental Figure 19: We conducted case-control comparisons for dementia-related neurodegenerative diseases. We also calculated the proportion of explained variance of overall cortical and sub-cortical structures regarding differences between neurodegenerative patients and healthy controls, as quantified by Partial $\eta^2$ values. a Partial $\eta^2$ values represent the proportion of explained variance of different cortical measurements for disease differences. Light purple indicates the 3T cortical regions, while dark purple represents P7T cortical regions. b Partial $\eta^2$ values represent the proportion of explained variance of different sub-cortical measurements for disease differences. Light blue indicates the 3T sub-cortical structures, while dark purple represents P7T sub-cortical regions.
